# Single-Cell Profiling Reveals Targetable Malignant T-Cell Subtypes and Immune Evasion Pathways in Sézary Syndrome

**DOI:** 10.1101/2025.05.20.25327833

**Authors:** Beth A. Childs, Eslam A. Elghonaimy, Praveen Ramakrishnan Geethakumari, Kiran A. Kumar, Joseph F. Merola, Heather W. Goff, Todd A. Aguilera

## Abstract

**Background:** Sézary syndrome (SS) is a rare and aggressive leukemic variant of cutaneous T-cell lymphoma (CTCL) with limited therapeutic options and a median survival of fewer than five years. Despite advances in how single-cell RNA sequencing (scRNA-seq) has improved the understanding and treatment of other cancer types, such insights in SS remain limited. The drivers of disease progression and immunologic dysfunction are incompletely defined, underscoring the need to characterize both malignant T cells (MTCs) and their interactions with surrounding immune populations.

**Objectives:** To systematically characterize malignant and non-malignant immune cells in CTCL, identify distinct malignant T-cell subtypes, and uncover transcriptional programs and immune evasion pathways with therapeutic relevance.

**Methods:** We analyzed scRNA-seq data on peripheral blood mononuclear cells (PBMCs) from 22 SS patients and 7 healthy controls. MTC subtypes were identified using a combination of transcriptional profiling, copy number variation (CNV) analysis, and T-cell receptor (TCR) clonotyping. CITE-seq was utilized on a subset of samples to correlate genetic findings with surface protein expression.

**Results:** We identified three MTC subtypes: (1) MTC central memory (CM), a Th2-skewed CM phenotype that constituted the dominant malignant population; (2) MTC effector/effector-memory (E/EM), a subset enriched in Th1-associated genes; and (3) MTC regulatory (Reg), a regulatory-like, exhausted phenotype, along with shared and subtype-specific gene signatures. The predominance of MTC CM suggests a stable malignant state, while relative rarity of MTC E/EM and MTC Reg may reflect treatment effects or disease progression. In addition to *KIR3DL2*, we identified *KIR2DL3* and *KIR3DL1* as upregulated immune-evasive receptors on MTCs and surrounding cells. Tensor factorization of ligand-receptor interactions revealed pro-inflammatory yet immunosuppressive signaling in myeloid cells converging on STAT3 activation.

**Conclusions:** This study defines three transcriptionally and functionally distinct MTC subtypes in SS, highlighting subtype-specific vulnerabilities that may inform personalized treatment strategies. Our findings suggest that targeting not only MTCs but also KIR-family receptor signaling and JAK-STAT activation in the immune microenvironment may have therapeutic implications. The identification of novel immunosuppressive pathways and cell survival mechanisms opens avenues for tailored interventions – including widespread KIR inhibition, repurposing JAK inhibitors, and other novel therapies – to improve patient outcomes in SS.

**Contributor Statement:** B.A. Childs conducted the conceptualization (equal), data curation (equal), formal analysis (lead), investigation (lead), methodology (equal), validation (equal), visualization (equal), writing – original draft (lead), writing – review & editing (supporting). E. Elghonaimy provided conceptualization (equal), data curation (equal), formal analysis (equal), investigation (equal), methodology (equal), resources (equal), supervision (equal), validation (equal), visualization (equal), writing – original draft (supporting), and writing – review & editing (supporting). P. R. Geethakumari provided data curation (supporting), investigation (supporting), resources (supporting), writing – review & editing (supporting). K. Kumar contributed conceptualization (supporting), data curation (supporting), resources (supporting), and writing – review & editing (supporting). J. F. Merola provided conceptualization (supporting), investigation (supporting), resources (supporting), writing – review & editing (supporting). H.W. Goff contributed conceptualization (equal), data curation (equal), formal analysis (supporting), funding acquisition (lead), investigation (supporting), methodology (supporting), resources (supporting), supervision (equal), visualization (equal), writing – review & editing (equal). T. A. Aguilera provided conceptualization of the project (equal), data curation (equal), formal analysis (equal), funding acquisition (equal), investigation (equal), methodology (equal), project administration (equal), resources (lead), supervision (lead), visualization (supporting), writing – original draft (supporting), writing – review & editing (equal).

T.A. Aguilera is the guarantor and accepts full responsibility for the overall content, had access to all the data, and controlled the decision to publish.

**Bulleted Statements:** *What is already known about this topic?:*

- Sézary syndrome is a rare, aggressive CTCL subtype associated with poor survival; a minority of patients achieve long-term remission with allogeneic stem cell transplant.
- *KIR3DL2* is an investigational target with ongoing trials (e.g., lacutamab, anti-*KIR3DL2* agent).
- JAK inhibitors have shown mixed effects, with anecdotal benefit and potential harm.
- The heterogeneous biology of Sezary cells remains poorly characterized, limiting diagnostic and therapeutic advancement.

*What does this study add?:*

- We define three distinct malignant T-cell subtypes in Sézary syndrome, each with clinical and therapeutic implications, and a unifying genetic signature.
- In addition to *KIR3DL2*, a known CTCL marker, we identify *KIR2DL3* and *KIR3DL1*, inhibitory receptors implicated in viral and autoimmune regulation, as additional immune evasion mechanisms.
- JAK/STAT activation in neighboring myeloid cells may be driven by Sezary cell signaling through *S100A8*, *S100A9*, *CD74*, *IL-10*, and *TNF*, a potentially targetable axis.

*What is the translational message?:*

- The predominance of a central memory–like malignant phenotype, alongside other distinct subsets reveal an opportunity for subtype-guided treatment strategies.
- Optimal therapeutic targeting may include both malignant T cells and interacting immune cells.
- Our findings identify targets that may be actionable, including KIR-family receptors and STAT3-related signaling.
- Insights from other T-cell disorders may guide repurposing of existing therapies to improve outcomes.

**Lay Summary: How Sezay Syndrome, a Type of Skin Lymphoma, Impacts the Immune System:** Sézary syndrome (SS) is a rare and aggressive form of skin cancer that starts in the blood. It is a type of cutaneous T-cell lymphoma (CTCL), where cancerous T cells (a kind of white blood cell) spread from the bloodstream to the skin. SS mainly affects adults and has limited treatment options, with most people living less than five years after diagnosis.

This study, based in the United States, aimed to understand how cancerous T cells behave in SS and how they interact with other immune cells. We used a method called single-cell RNA sequencing to examine over 100,000 individual immune cells from the blood of 22 patients with SS and 7 healthy individuals. This allowed us to see which genes were active in each cell.

We found that the cancerous T cells in SS are not all the same. Instead, they fall into three distinct groups: one resembled central memory T cells that typically live in the blood, one looked more aggressive and inflammatory, and one resembled exhausted regulatory immune cells. We also discovered that cancer cells and surrounding immune cells expressed molecules called KIRs, which may help them avoid being attacked. Other immune cells released signals that turned on a pathway called JAK-STAT, which may further protect the cancer.

These findings reveal new ways that SS protects itself and avoids the immune system. Understanding these escape routes may help guide future treatments, including drug combinations that block these signals and improve patient outcomes.

**Abbreviated Abstract:** Single-cell RNA and protein profiling of Sézary syndrome reveals three distinct malignant T-cell subtypes and uncovers convergent immunosuppressive signaling through KIR-family receptors and STAT3 activation. These findings expose targetable pathways driving immune evasion and offer a foundation for more personalized therapeutic strategies in this aggressive form of cutaneous T-cell lymphoma.

## Introduction

Sézary syndrome (SS) and leukemic variants of mycosis fungoides (MF) represent highly aggressive subtypes of cutaneous T-cell lymphoma (CTCL), a rare non-Hodgkin lymphoma characterized by clonal expansion of skin-homing T cells. These leukemic CTCLs are associated with rapid progression, immune dysfunction, and poor clinical outcomes, with median survival dramatically decreasing to 1–4 years upon significant peripheral blood involvement.^1^ Despite recent advancements in diagnostic technologies such as flow cytometry-based immunophenotyping and molecular T-cell receptor (TCR) clonality analyses, definitive classification remains elusive. Current immunophenotypic markers—including CD4+CD26−, CD4+CD7− subsets, and TCR Vβ expansions—lack the sensitivity and specificity required for accurate disease stratification.^2^

The pathogenesis of leukemic CTCL remains incompletely understood, underscoring both diagnostic uncertainty and biological complexity. Fundamental questions persist regarding the precise cellular origins, transcriptional programs, and immune evasion mechanisms that drive disease progression. Addressing these knowledge gaps is critical for the development of effective targeted therapies.^3,4^

To overcome these challenges and identify malignant cells and their physiological state, we employed a high-resolution approach based on single cell RNA sequencing (scRNA-seq) that enabled systematic characterization of the tumor and peripheral immune landscape. This work characterizes previously unrecognized transcriptional subsets among malignant T cells (MTCs) and nominates mechanisms of immune evasion. We evaluated peripheral blood mononuclear cells (PBMCs) from 22 leukemic CTCL patients and 7 healthy controls (HCs), then integrated these data with paired TCR sequencing and CITE-seq proteomic profiling.^2,5-7^ Our results nominate distinct immune evasion strategies in CTCL, including upregulation of inhibitory killer immunoglobulin like receptors (KIRs) beyond *KIR3DL2* and STAT3-driven ligand-receptor signaling through *S100A8/A9* and *CD74*. Further, certain vulnerabilities appear amplified in specific MTC subsets. Altogether, these findings propose novel mechanisms of immune evasion and reveal both subset-specific and broader therapeutic opportunities that could help improve patient outcomes.

## Methods

Details explaining patient recruitment; sample collection, compilation, and processing; droplet-based scRNA-seq; data analysis workflows; secondary analyses; and methodologies for differential expression, cell-cell communication, and gene set enrichment analyses are provided in Appendix S1.

## Results

### Project design and cell annotations

To characterize the immune landscape of CTCL at single-cell resolution, we analyzed scRNA-seq data from 144,321 PBMCs, including 101,710 cells from 22 patients with SS or leukemic MF (8 from our institution and 14 from public datasets) and 42,611 cells from seven publicly available HCs (144,321 cells total) (Figure 1A).^5-9^ The mean age of patients with CTCL was 67 years, and the cohort included 12 women and 10 men (Table 1). Following sample- and cell-level quality control and data integration, we retained 70,519 high-quality cells from CTCL and 37,100 from HCs for downstream analysis (Supplementary Figure 1A–C). Unsupervised clustering revealed major immune lineages, including T cells, myeloid cells, NK cells, and B cells. Further subclustering identified canonical T cell subsets as well as diverse monocyte and dendritic cell (DC) populations, naïve B cells, and NK cells (Figure 1B; Supplementary Figure 1D–G; Tables S1-3).^10-12^

**Figure 1:**
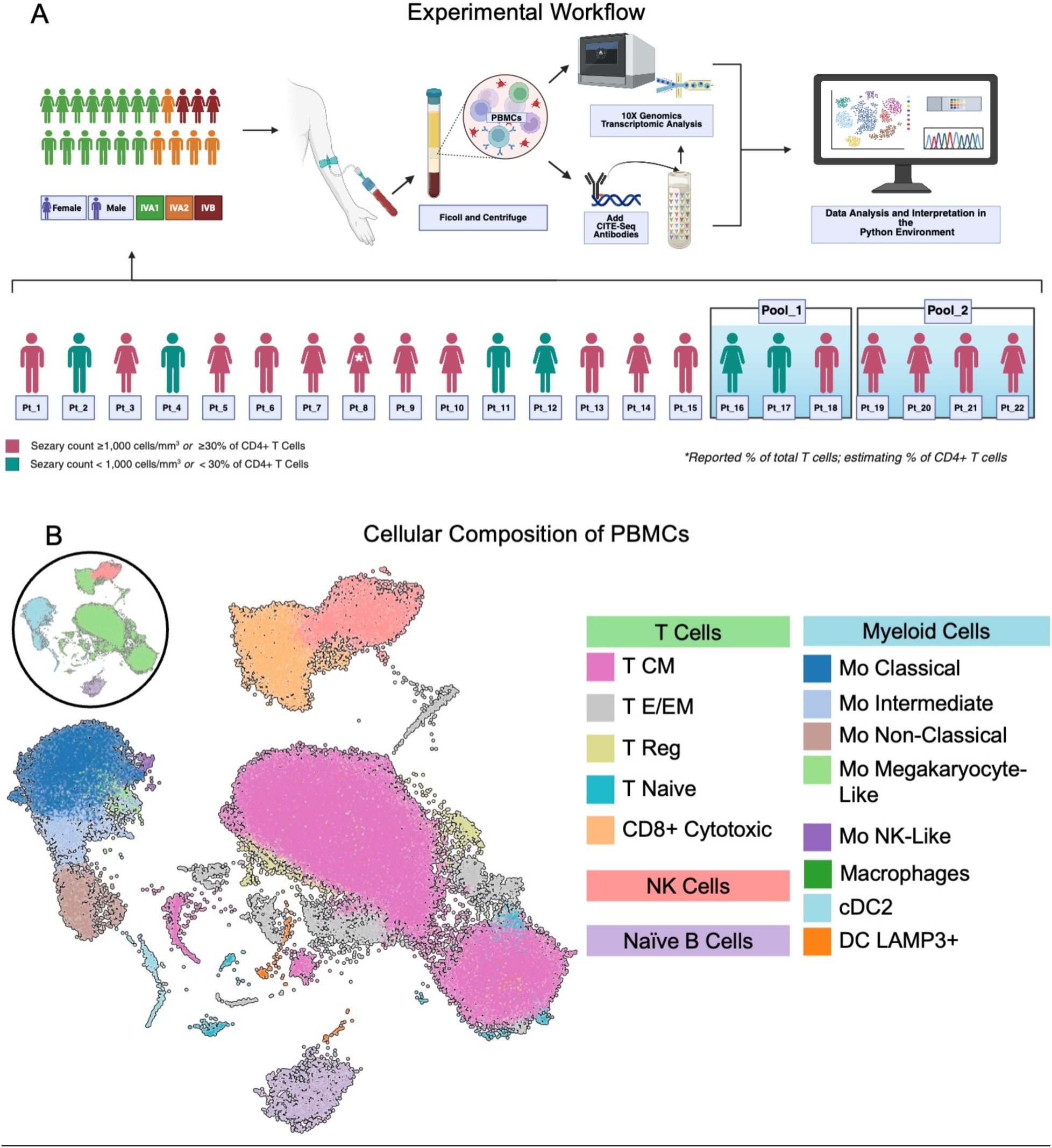
Project workflow and cellular composition in CTCL. (a) Overview of single-cell RNA sequencing (scRNA-seq) analysis pipeline, encompassing preprocessing, quality control, and Harmony-based integration of 101,710 peripheral blood mononuclear cells (PBMCs) from 22 patients with cutaneous T-cell lymphoma (CTCL). Downstream analyses included 70,519 high-quality cells. (b) UMAP visualization of all PBMCs, colored by major immune lineages and refined subsets. Broad populations include T cells, natural killer (NK) cells, naïve B cells, and myeloid cells. Further subclustering identified distinct T cell phenotypes (e.g., central memory [CM], effector/memory [E/EM], regulatory [Reg], naïve, and cytotoxic) and diverse myeloid subsets, including classical, intermediate, and non-classical monocytes (Mo), Mo megakaryocyte-like, Mo NK-like, type 2 conventional dendritic cells (cDC2s), macrophages (MP), and LAMP3+ dendritic cells (DCs).

**Table 1:** Patient Demographics and Clinical Characteristics. This table summarizes the demographics and clinical characteristics of the patient cohort analyzed in this study. It includes the sample identifiers grouped by study pool, clinical diagnoses, patient age in years, sex, Sézary cell count expressed as absolute cells per cubic millimeter or as a percentage of total CD4+ T cells (*percentage provided as total T cells in study GSE165623), prior treatments, and treatments administered within two weeks of sample collection (only systemic and lymphoma-specific therapies listed). The final column provides a brief clinical description for patients diagnosed with cutaneous T-cell lymphoma (CTCL) or lists other known diagnoses for healthy control (HC) participants. ***Abbreviations:*** *BSA: Body surface area; CysA: Cyclosporine A; DVT: Deep vein thrombosis; ECP: Extracorporeal photopheresis; HC: Healthy control; HLD: Hyperlipidemia; IFNa: Interferon-alpha; MMF: Mycophenolate mofetil; MTX: Methotrexate; NA: Not available; NBUVB: Narrowband ultraviolet B phototherapy; Pred: Prednisone; TBSA: Total body surface area; TSEB: Total skin electron beam radiation therapy*

| Pool/<br>Group | Sample | Age<br>(Years) | Sex | Sezary<br>Count<br>(cells/mm<sup>3</sup>)/<br>Percentage of CD4+<br>T Cells | Prior<br>Treatments | Treatment at<br>Time of<br>Sampling | Clinical<br>Description |
| --- | --- | --- | --- | --- | --- | --- | --- |
|  | Pt_1 | 76-80 | M | 31% | Mogamulizumab | NBUVB, Romidepsin | Erythroderma, >90% BSA with patches and plaques |
|  | Pt_2 | 61-65 | M | 893 | Bexarotene, IFNa | Vorinostat, ECP | Not available. |
|  | Pt_3 | 71-75 | F | 6,129 | MTX | Bexarotene | Not available. |
|  | Pt_4 | 66-70 | M | 110 | NA | IFNa, ECP | Not available. |
|  | Pt_5 | 81-85 | F | 1,968 | NBUVB | ECP | Not available. |
|  | Pt_6 | 71-75 | M | 1,200 | Pred | ECP, bexarotene | Not available. |
|  | Pt_7 | 86-90 | F | 6,300 | NA | Romidepsin | Not available. |
|  | Pt_8* | 61-65 | F | 13%* | Bexarotene, TSEB, gemcitabine | 7-year no treatment | Generalized patches, plaques, and tumors |
|  | Pt_9 | 36-40 | F | 36% | MTX, IFNa, Romidepsin | None for 2 weeks | Plaques |
|  | Pt_10 | 46-50 | F | 60% | NA | NA | <1% TBSA patches |
|  | Pt_11 | 61-65 | M | 17% | NA | NA | Erythroderma |
|  | Pt_12 | 71-75 | F | 21% | NA | NA | Plaques/rashes |
|  | Pt_13 | 56-60 | M | 36% | NA | NA | Erythroderma |
|  | Pt_14 | 71-75 | F | 60% | NA | ECP | 85% TBSA, erythroderma |
|  | Pt_15 | 76-80 | M | 46% | NA | NA | 100% TBSA, erythroderma |
| Pool_1 | Pt_16 | 76-80 | F | 9% | ECP, IFNa | IFNa | Patches and plaques |
|  | Pt_17 | 51-55 | M | 67% | CysA | Romidepsin | Erythroderma, keratoderma |
|  | Pt_18 | 51-55 | M | 92% | MMF, adalimumab, ECP | IFNa | Erythroderma, keratoderma |
| Pool_2 | Pt_19 | 76-80 | F | 85% | NA | NA | Erythroderma, keratoderma |
|  | Pt_20 | 71-75 | F | 73% | NA | Acitretin | Patches and thin plaques on trunk, buttocks, and extremities |
|  | **Pt_21** | 41-45 | M | 76% | Acitretin,<br>INFa, ECP,<br>Romidepsin | Bexarotene | Erythroderma,<br>keratoderma |
|  | **Pt_22** | 76-80 | F | 98% | IFN, ECP,<br>Bexarotene,<br>Acitretin,<br>NBUVB | Romidepsin | Erythroderma |
| **Healthy**<br>**Control** | **HC_1** | 56-60 | F | NA | NA | NA | HLD,<br>bipolar/depression |
|  | **HC_2** | 61-65 | F | NA | NA | NA | DVT, depression,<br>recent flu |
|  | **HC_3** | 66-70 | F | NA | NA | NA | Schizophrenia,<br>seizures |
|  | **HC_4** | 56-60 | M | NA | NA | NA | Asthma |
|  | **HC_5** | 31-35 | M | NA | NA | NA | NA |
|  | **HC_6** | 56-60 | M | NA | NA | NA | HLD, lung disease |
|  | **HC_7** | 41-45 | M | NA | NA | NA | NA |

### Heterogeneity of malignant T cells in CTCL reveals three dominant subtypes

Among patients with CTCL, MTCs were identified using a multifactorial approach that integrated dominant TCR clonotypes (when available), inferred CNVs, and transcriptional divergence from HCs. Clusters with >50% of clonally expanded cells were considered malignant, consistent with prior literature (Appendix S1, Figure 2A, Supplementary Figure 2A).^5,6^ No dominant CDR3 sequences were shared across samples. Using these criteria, we identified 34,974 MTCs and 18,094 non-malignant T cells in CTCL patients.

**Figure 2:**
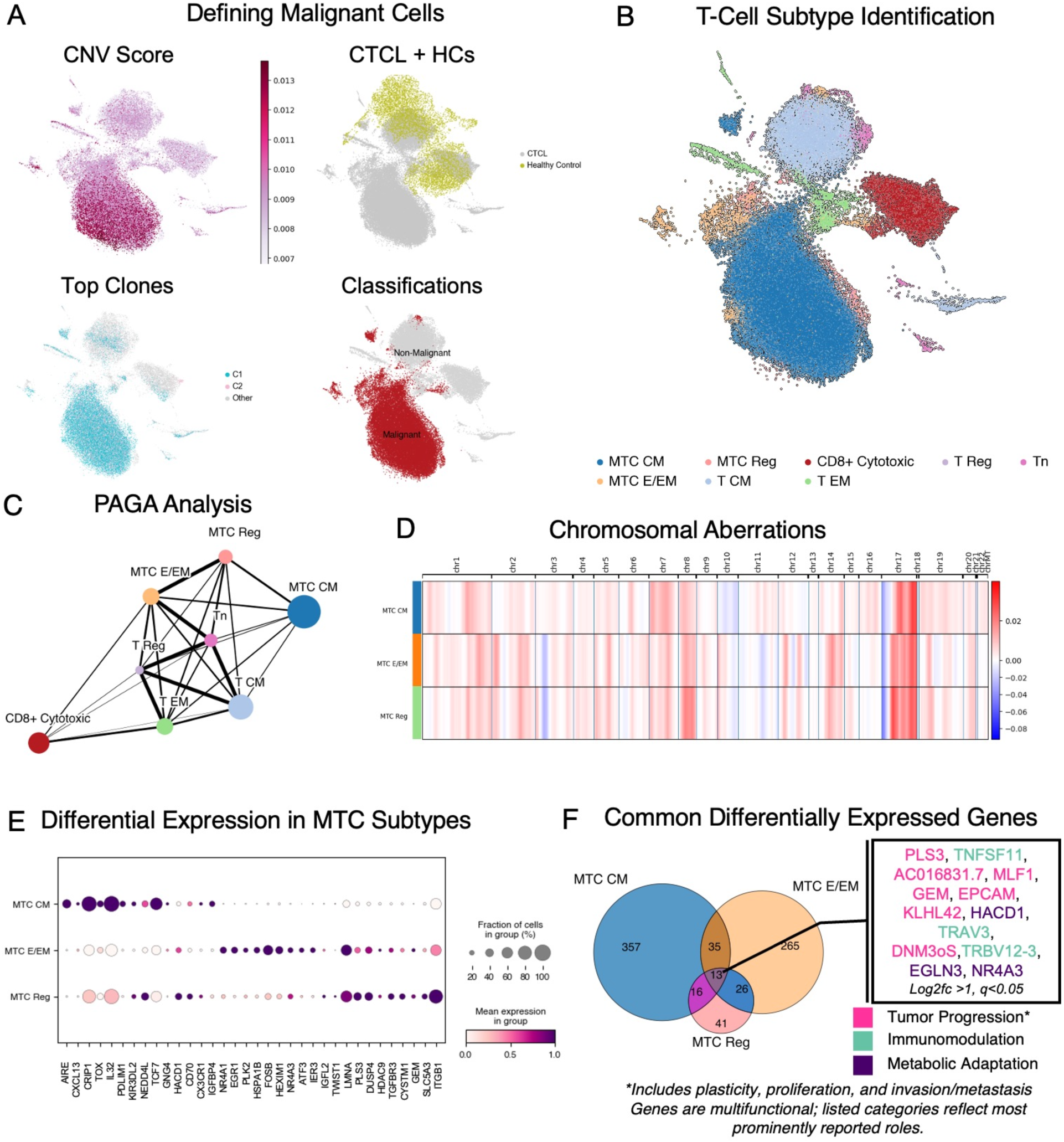
Diverse malignant T cell subtypes reveal distinct pathogenic and therapeutic signatures. (a) Panels illustrating malignancy criteria including copy number variation (CNV) scores, comparative visualization with healthy controls, malignant versus benign T-cell labeling, and top clonotype frequency based on T-cell receptor (TCR) sequencing data. (b) UMAP visualization of T-cell clusters in CTCL patients, identifying distinct malignant T-cell (MTC) subtypes—MTC CM (central memory-like, Th2/Th22-skewed), MTC E/EM (effector/effector memory, Th1-biased), and MTC Reg (regulatory/exhausted-like). (c) Plot illustrating connectivity and potential transitional relationships between malignant and benign T-cell subsets, suggesting possible plasticity between MTC E/EM and MTC Reg. (d) Heatmap summarizing distinct CNV patterns across malignant T-cell subtypes, generated with inferCNVpy. (e) Dot plot showing the top differentially expressed genes distinguishing each malignant T-cell subtype from their benign counterparts (e.g., MTC CM vs. T CM). (f) Venn diagram displaying shared significantly upregulated genes (log₂ fold change [logFC] > 1, q < 0.05) across the three MTC subtypes.

Unsupervised clustering of T cells from CTCL patients revealed three transcriptionally and clonally distinct malignant subsets: MTC CM (central memory-like), MTC E/EM (effector/effector memory-like), and MTC Reg (regulatory/exhausted-like) (Figure 2B). Subsets were distinguished using characteristic transcriptional profiles (Appendix S1). We next used CITE-seq profiling (n = 7), a proteomic assessment using oligo labeled antibodies for single-cell sequencing, which validated transcriptomic annotations at the protein level. This confirmed increased CCR4 and reduced CD5 and CD7 expression in MTCs relative to benign T cells.

CD25 expression was restricted to MTC Reg, while PD-1 was observed on a minority of MTC CM cells (Supplementary Figure 2B). Partition-based Graph Abstraction (PAGA) trajectory inference suggested that MTC CM forms a transcriptionally distinct and potentially terminal state, whereas MTC E/EM and MTC Reg exhibited closer connectivity and possible lineage plasticity (Figure 2C).

Chromosomal CNV analysis revealed distinct patterns across malignant subsets: MTC CM and MTC Reg showed strong, recurrent alterations on chromosomes 7, 8, and 17, while MTC E/EM exhibited a more diffuse pattern with modest CNVs across multiple chromosomes, including 8, 12, 14, and 17 (Figure 2D). Subtype proportions varied by patient, with MTC CM predominant in most individuals, while MTC E/EM and MTC Reg were present in select cases (Supplementary Figure 2C).

### Functional signatures of malignant T cells provide insight into biologic activity

To isolate transcriptional features specific to malignancy rather than baseline cell phenotype, we performed differential expression analysis by comparing each malignant subtype to its closest benign counterpart: MTC CM vs. T CM, MTC E/EM vs. T EM, and MTC Reg vs. T Reg (Figure 2E; Supplementary Figure 2D; Tables S4-6). This approach revealed distinct sets of dysregulated genes characterizing each malignant state.

MTC CM exhibited a Th2/Th22-skewed, immune-evasive profile marked by upregulation of genes previously linked to CTCL such as *AIRE, CXCL13, TCF7, KIR3DL2*, and *NEDD4L*, alongside decreased *CD7*.^6,8,13-15^ *CRIP1*, associated with immunosuppression and poor prognosis in other cancer types, and *GNG4*, implicated in the progression of other malignancies, were newly identified transcripts in SS cells (all q<0.0001).^16,17^ While many of these markers have been individually reported, our analysis reinforces their relevance within a cohesive malignant CD4+ T-cell phenotype.

The less common subsets, MTC E/EM and MTC Reg, also exhibited distinct transcriptional characteristics. MTC E/EM demonstrated a stress-adaptive, metabolically reprogrammed state with high expression of *FOSB, EGR1, HSPA1B, NR4A1*, and *LMNA*.^18-21^ MTC Reg overlapped partially with both MTC CM and MTC E/EM, expressing shared immune-evasive and stem-like transcripts (e.g., *KIR3DL2, NEDD4L, CD70*) as well as survival-associated genes enriched in MTC E/EM (e.g., *LMNA, DUSP4*).^13,22-24^

Despite this heterogeneity, we identified a set of 13 genes consistently upregulated across all malignant subsets, representing a conserved malignant profile enriched for transcripts associated with immunomodulation, proliferation, invasion, and metabolic adaptation, including *PLS3*, *MLF1*, *GEM*, *DNMO3S*, *HACD1*, and *NR4A3* (Figure 2F). ^25-35^

Clinically, MTC E/EM cells were more frequently observed in untreated patients (e.g., Pt_10– 15), while MTC Reg cells were enriched in those receiving HDAC inhibitors (HDACis) (e.g., Pt_2, Pt_7, Pt_8), consistent with prior associations between *FOXP3* expression and HDACi exposure.^5^ However, these trends were not uniform, and dataset limitations restricted our ability to capture individual patterns (Supplementary Figure 2C; Table 1), underscoring both biological and clinical heterogeneity.

### Altered nonmalignant cell composition and gene expression in CTCL

While initial annotations (Figure 1) established the peripheral immune cell types present in CTCL, we next sought to define how these compartments differ from healthy individuals to better understand disease-associated remodeling of the non–T-cell landscape. Differential gene expression analysis revealed widespread transcriptional alterations across B cells, NK cells, monocytes, and DCs in CTCL compared to healthy controls (Figure 3A). Cell type assignments were supported by canonical marker expression (Figure 3B) and further validated using CITE-seq surface protein profiling (n=7; Supplementary Figure 3A).

**Figure 3:**
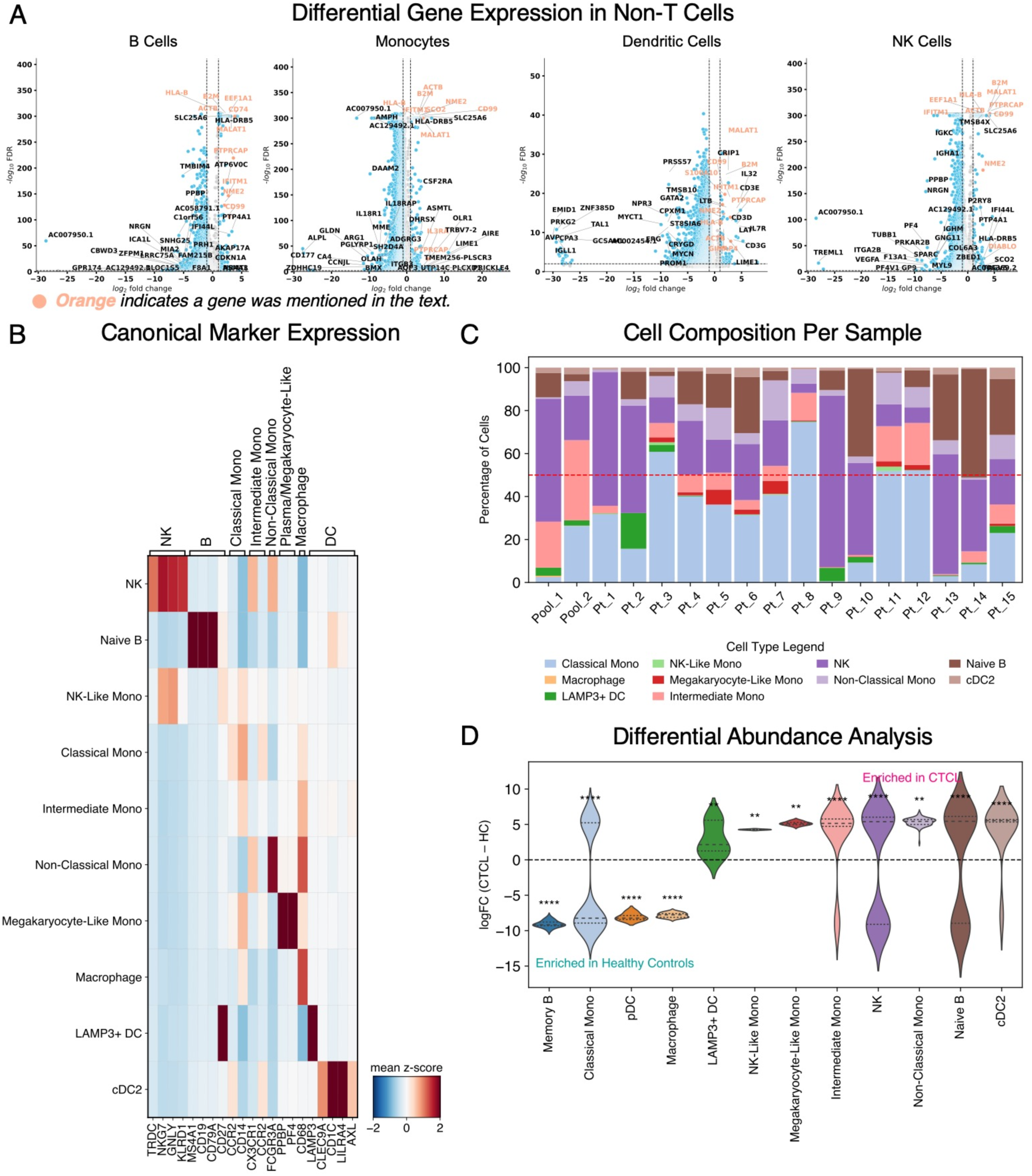
Non-T cells demonstrate distinct profiles in patients with CTCL compared to healthy controls. (a) Volcano plots showing differentially expressed genes (DEGs) between CTCL patients and healthy controls in NK cells, DCs, monocytes (Mono), and B cells. Axes represent log2 fold change and –log10 false discovery rate (FDR). (b) Heatmap displaying canonical marker expression across non–T cell subsets within CTCL patient samples. (c) Bar plot depicting the proportional abundance of non–T cell populations per sample. (d) Violin plots comparing cell neighborhood enrichment scores between CTCL patients and healthy controls, indicating microenvironmental alterations in non–T cell spatial context. Statistical significance is annotated as: * = p ≤ 0.05, ** = p ≤ 0.01, *** = p ≤ 0.001, **** = p ≤ 0.0001. *Abbreviations: pDC, plasmacytoid dendritic cell*.

We identified 25 genes significantly upregulated across all four non–T cell types, including *MALAT1*, *IL32*, *CD99*, *NME2*, *HLA-B*, *IFITM1*, *B2M*, *ACTB*, and *PTPRCAP*, suggesting convergent activation, stress adaptation, and migratory signaling.^36-51^ Multiple mitochondrial genes were also upregulated, indicating metabolic adaptation (Figure 3A, Supplementary Figure 3B).

Cell type–specific changes added further resolution to this dysregulated immune landscape. B cells showed increased *CD74* and *EEF1A1* expression. *CD74* facilitates antigen presentation and promotes tumor-supportive signaling in other cancer types, and *EEF1A1* is involved in protein synthesis and cellular stress responses (Table S7).^52,53^ Monocytes upregulated *SCO2* and *IL3RA*. *SCO2* regulates mitochondrial metabolism in response to stress, and *IL3RA* indicates chemokine-driven recruitment and immune modulation (Table S8).^40,54,55^ DCs upregulated *S100A10* and *GIMAP4*, suggesting roles in immune regulation, infiltration, and survival (Table S9).^56,57^ NK cells upregulated *EEF1A1,* shared with monocytes and B cells, and *DIABLO*, a pro-apoptotic gene (Figure 3A, Table S10).^58^

Although cell-type frequencies varied across individual samples, as previously reported in CTCL, proportional analysis revealed significant and consistent differences after adjusting for sample-level variation (Figure 3C).^5,7^ Non-classical and intermediate monocytes, LAMP3+ DCs, and naïve B cells were expanded in CTCL, while classical monocytes, plasmacytoid DCs, and mature B cells were more abundant in healthy controls (Figure 3D; Supplementary Figure 3C– D). These shifts suggest a reorganization of the non-T-cell compartment, likely driven by chronic inflammation and tumor-mediated immune reprogramming.

### Malignant T cells exploit KIR-family receptors to evade immune surveillance

To contextualize immune interactions among MTCs and immune cells, we examined the cell– cell communication landscape. Supplementary Figure 4A shows the mean ligand-receptor interaction strengths across cell types, highlighting Sézary cells as central participants in immune signaling networks, particularly with NK cells, NK-like monocytes, CD8+ T cells, and naïve B cells. This is notable because Sézary cells may either be suppressed by surrounding immune cells or, conversely, co-opt inhibitory signaling pathways to promote their own survival or self-renewal.

While analyzing ligand–receptor interactions, we observed enhanced signaling involving the inhibitory *KIR3DL2*, *KIR3DL1*, and *KIR2DL3* receptor genes in MTCs.^59^ This prompted a deeper investigation of the KIR axis, given its established role in immune regulation and its emerging relevance in CTCL.^13^ *KIR3DL2* (CD158k) is known to be overexpressed in CTCL biopsy samples, supporting the rationale for a first-in-class anti-KIR3DL2 antibody.^13,60^ In our dataset, we observed upregulation of *KIR3DL2*, *KIR3DL1*, and *KIR2DL3*, which may contribute to MTC survival.

The most significantly elevated interaction was Major Histocompatibility Complex I (*MHCI)* (combined *HLA-A, HLA-B, HLA-C,* and *B2M*)*–KIR3DL2* between MTCs and surrounding cells, especially in MTC CM and MTC Reg when compared to benign counterparts (Figure 4A; Supplementary Figure 4B). Elevated signaling was also observed for *KIR3DL1* and *KIR2DL3* and extended beyond traditional NK cells to involve CD8+ T cells, monocytes, and B cells (Supplementary Figures C-D). These findings were consistent with differential gene expression analysis, which revealed significant upregulation of *KIR3DL2*, *KIR2DL3*, and *KIR3DL1* in MTC CM versus T CM, and of *KIR3DL2* in MTC Reg versus T Reg (Figure 4B).

**Figure 4:**
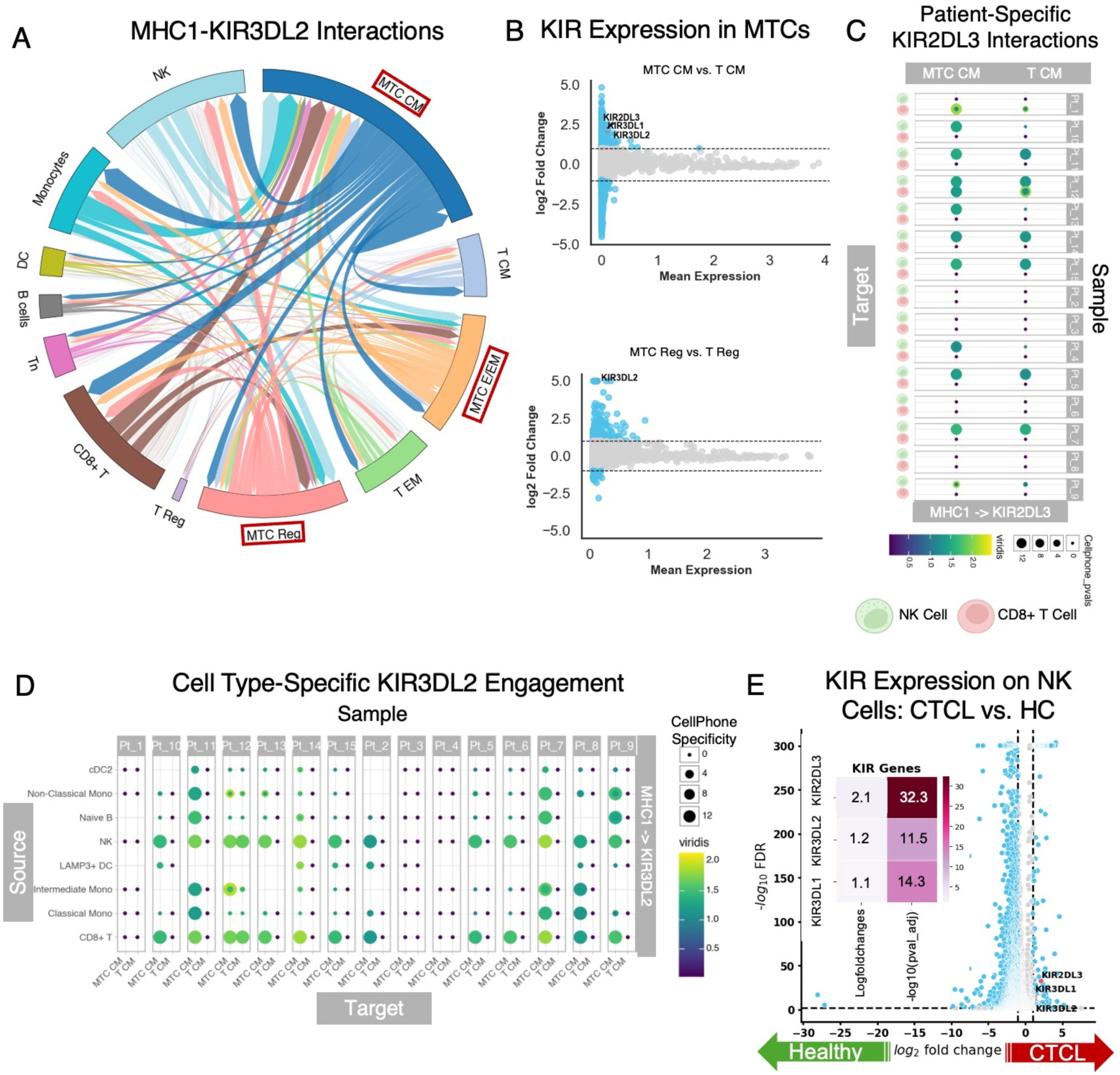
Malignant T CM and T Reg Cells Engage in Bidirectional Immunosuppressive Interactions via KIR-Family Receptors. (a) Circos plot showing increased major histocompatibility complex class I (MHCI)–Killer Cell Immunoglobulin Like Receptor (KIR) 3DL2 interactions between MTC CM and MTC Reg and surrounding immune cell populations. (b) Mean average (MA) plot comparing expression of KIR-family receptors in MTC CM versus benign T CM, highlighting significant upregulation in malignant cells. (c) Dot plot illustrating patient-specific signaling from MTC CM to NK and Cytotoxic T cells (CD8+ T) via KIR2DL3. (d) Dot plot showing enhanced MHC class I–KIR3DL2 engagement in MTC CM compared to benign T CM, reinforcing its role as a potential therapeutic target. (e) Volcano plot and heatmap showing increased expression of KIR2DL3, KIR3DL1, and KIR3DL2 in NK cells from CTCL patients.

To better understand these interactions at the patient level, we examined source–target cell-type signaling patterns. MTC CM consistently demonstrated stronger outgoing *KIR2DL3*-mediated interactions with NK and CD8+ T cells compared to benign T CM in most patients (Figure 4C). Additionally, across patients, MTCs received more frequent and stronger *KIR3DL2* input from diverse immune cell types including NK cells, CD8+ T cells, and monocytes compared to their benign counterparts (Figure 4D), supporting a model of widespread bidirectional immune suppression. This pattern of enhanced KIR signaling was further supported by analysis of the broader immune environment, where all three KIR-family genes of interest were significantly upregulated in NK cells from CTCL patients versus HCs (Figure 4E).

### Myeloid JAK/STAT signaling suggests IL-10/STAT3-mediated support of malignant T cells

To understand how the tumor microenvironment contributes to immune dysfunction in CTCL, we modeled cell–cell communication among non–T cell populations from CTCL patients and HCs using ligand–receptor interaction data. To identify overarching patterns in these interactions, we applied tensor factorization, a dimensionality reduction technique that captures coordinated variation across multiple axes of complex data.^61^ In this context, tensor factorization decomposed a multi-dimensional matrix of ligand–receptor interactions across cell types and patients, revealing latent communication signatures that differentiate disease states. This analysis uncovered several dysregulated signaling programs, with Factors 2 and 5 emerging as the most distinct between CTCL and HC samples (Figure 5A–B, Supplementary Figure 5A–B).

**Figure 5:**
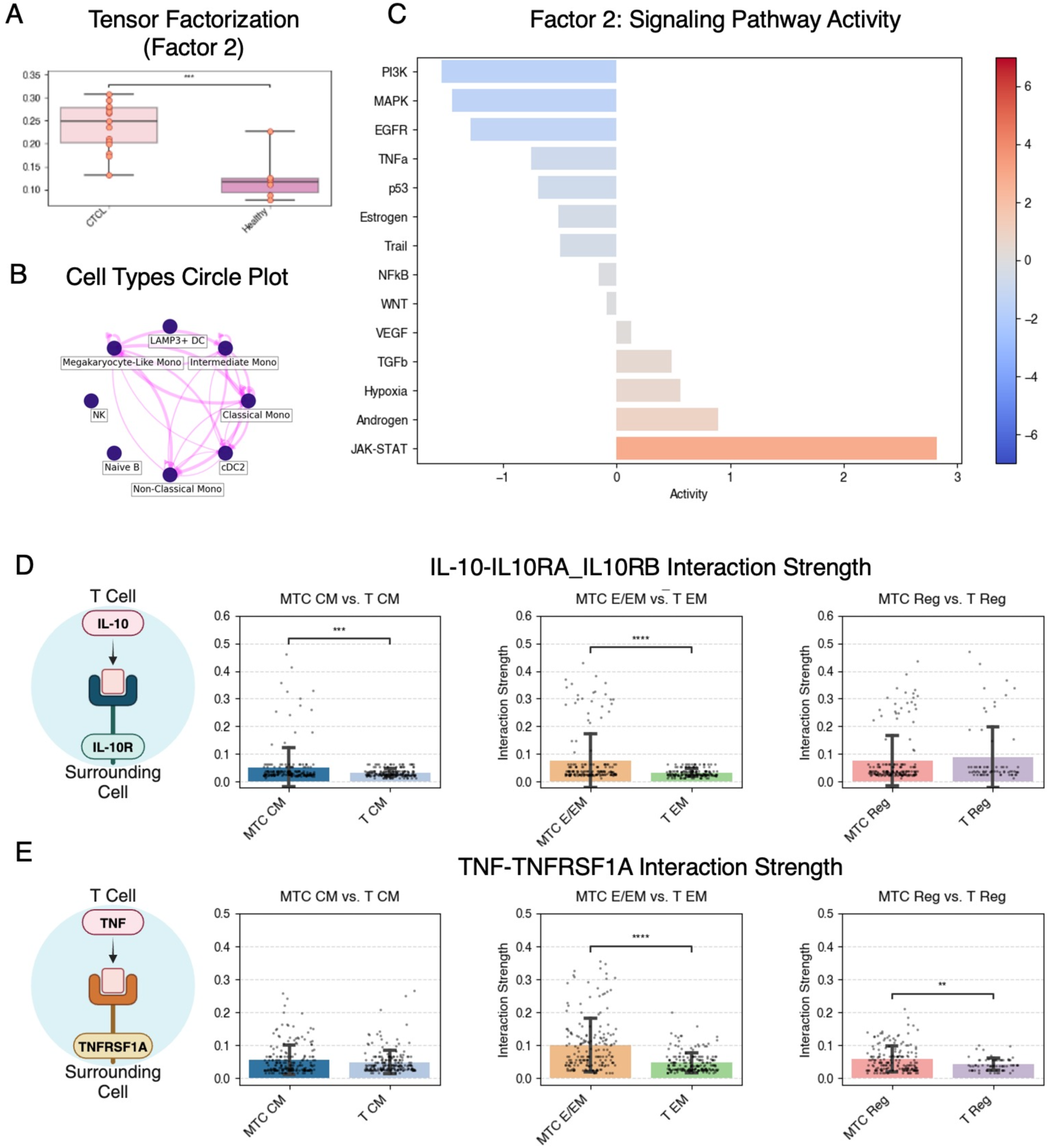
JAK/STAT activation in surrounding myeloid cells suggests targetable signaling pathways. (a) Boxplot showing Factor 2 scores from tensor decomposition analysis (LIANA), demonstrating significantly higher signaling in CTCL patients compared to healthy controls (q < 0.01). Statistical significance is annotated as: * = p ≤ 0.05, ** = p ≤ 0.01, *** = p ≤ 0.001, **** = p ≤ 0.0001. (b) Circle plot highlighting the primary cell types contributing to Factor 2, notably monocytes and dendritic cells. (c) Horizontal bar plot displaying elevated Janus kinase/signal transducer and activator of transcription (JAK-STAT) pathway activity discovered from Factor 2 in Monos and DCs from CTCL patients relative to healthy controls. (d) Boxplots illustrating significantly stronger interleukin (IL)-10→IL10RA_IL10RB interactions from MTCs (MTC CM and MTC E/EM) compared to benign subsets. (e) Boxplots demonstrating increased tumor necrosis factor (TNF)→TNF receptor superfamily member 1A (TNFRSF1A) signaling from MTC E/EM and MTC Reg to other immune cells, suggesting paracrine immunosuppressive effects. *Statistical significance is annotated as: * = p ≤ 0.05, ** = p ≤ 0.01, *** = p ≤ 0.001, **** = p ≤ 0.0001*.

Factor 2, significantly enriched in CTCL (q < 0.0001), captured a dominant signaling module involving *S100A8* and *S100A9* interactions with *CD68, CD36*, and *ITGB2* on myeloid cells (Supplementary Figure 5C). These calgranulin-mediated interactions, associated with chronic inflammation and myeloid activation, were predominantly active in monocytes and DCs, which exhibited nearly threefold higher JAK/STAT pathway activity compared to HCs (Figure 5C).

This transcriptional program aligns with a tumor-supportive myeloid phenotype observed in other cancers and may contribute to immune suppression via downstream STAT3 activation.^62-64^

Supporting this, MTCs exhibited significantly increased *IL-10–IL10RA_IL10RB* signaling toward surrounding immune cells (Figure 5D). As interleukin (IL)-10 is a potent activator of JAK1/TYK2–STAT3 signaling, this may represent a mechanism by which MTCs directly reprogram the microenvironment to favor immune tolerance and tumor persistence.^65^ Additionally, *TNF–TNFRSF1A* interactions were significantly elevated in MTC E/EM and MTC Reg subtypes (Figure 5E), further implicating pro-inflammatory tumor necrosis factor (TNF) signaling in the context of STAT3-mediated immune dysregulation.^66^ While this immunoregulatory axis could contribute to tumor persistence, its precise role in CTCL pathogenesis remains unclear, warranting further investigation.

By contrast, Factor 5, enriched in HCs (q < 0.0001), was defined by homeostatic immune interactions involving GPCR signaling (*GNAI2–S1PR4, GNAI2–ADCY7*), chemotaxis (*HMGB1– CXCR4*), and cell adhesion (*ADAM10–CD44*). *GNAI2* encodes Gαi2, a key negative regulator of adenylyl cyclase and modulator of multiple immune pathways. Loss of *GNAI2* expression has been linked to tumor progression in ovarian cancer and impaired T cell migration in primary immunodeficiency.^67,68^ Its attenuation in CTCL may contribute to aberrant immune cell positioning and unchecked T cell activation, further disrupting microenvironmental homeostasis.

Factors 4 and 6, both significantly enriched in CTCL, highlight CD74-centered immunoregulation as a recurring theme. Key interactions included *MIF–CD74–CXCR4, COPA– CD74*, and *APP–CD74*, all known to promote STAT3 activation and immune suppression.^69^ Factor 6 additionally featured *LILRB1–MHCI* interactions, suggesting checkpoint-like inhibition. Given broad upregulation of *CD74* across non–T cells in CTCL (log₂FC = 1.44, q < 0.0001), these findings position it as a central node of immune dysfunction and a promising therapeutic target.^70,71^ Notably, Factor 3 also showed elevated average *MHC–KIR* interaction strengths among non-T cells in CTCL compared to HCs, echoing the KIR-driven inhibitory signaling observed in MTCs, though this trend did not reach statistical significance.

## Discussion

SS is increasingly recognized as a clinically and transcriptionally heterogeneous disease.^72^ In this study, we mapped CTCL blood composition using multimodal sequencing, identifying three distinct MTC subtypes with potential therapeutic vulnerabilities: MTC CM (central memory-like, immune evasive, most prominent), MTC E/EM (effector/effector memory-like, metabolically adapted), and MTC Reg (regulatory/exhausted-like, immunosuppressive). We developed a transcriptional signature distinguishing malignant from benign T cells, highlighting consistent upregulation of genes such as *DNM3oS* and *PLS3* across subtypes, reinforcing their potential role in CTCL pathogenesis.^73^

Each MTC subtype suggests distinct vulnerabilities with therapeutic relevance. MTC CM, the most prevalent subtype, exhibited Th2/Th22 polarization and central memory markers (*CCR7, SELL, TCF7*), supporting the idea that central memory T cells are the archetypal Sézary cell.^74^ It also demonstrated high expression KIR-family receptors and *CXCL13*. Lacutamab (anti-*KIR3DL2*) and *CXCL13-*targeted therapies (under investigation for inflammatory bowel disease and rheumatoid arthritis) represent promising therapeutic avenues.^75,76^ MTC E/EM displayed NR4A family activation, suggesting susceptibility to NR4A-modulating agents such as GLP-1R agonists.^77^ MTC Reg showed elevated *ITGB1* expression, an investigational target in other cancer types, and modest enrichment in HDAC inhibitor-treated patients, suggesting treatment may influence phenotype.^5,78^ *EPCAM*, broadly upregulated across MTCs, may offer an additional therapeutic target, aligning with emerging EpCAM-directed CAR-T strategies in other malignancies.^79^

Transcriptional remodeling of non-T cell compartments may also serve a critical function in supporting Sézary cell adaptation and persistence, including the upregulation of *MALAT1,* which is induced by STAT3 and promotes IL-10 secretion and pro-tumor myeloid recruitment.^80-83^ IL-32, a cytokine implicated in other cancers and autoimmune diseases, was also elevated in myeloid cells.^84^ These findings suggest non–T cells may actively contribute to CTCL pathogenesis and represent underexplored targets for intervention.

Our data suggest the KIR-family receptors could be key mediators of immune evasion or support cell survival. Clinical trials of the anti-KIR3DL2 antibody lacutamab (IPH4102) demonstrated durable responses and improved quality of life in relapsed or refractory SS, with 89% of patients showing stable or improved disease and a favorable safety profile. Although 14% developed lymphopenia, *KIR3DL2*-expressing NK cell counts remained largely unaffected.^60^ In addition to *KIR3DL2*, we observed upregulation of *KIR2DL3* and *KIR3DL1* across MTCs and innate immune cells, suggesting a self-reinforcing inhibitory network suppressing both adaptive and innate responses.^13,60,85-87^ Enhanced *MHCI-KIR* interactions further support this model.

Finally, our analysis underscores JAK/STAT signaling as a potential central axis of immune dysfunction in CTCL. Tensor factorization revealed *S100A8, S100A9*, and *CD74*-mediated signaling converging on STAT3 activation, partly driven by MTC-derived *IL-10* and *TNF– TNFRSF1A*. *S100A9–TLR4* signaling, previously linked to NF-κB–driven tumor growth in CTCL, may be reversible with agents like tasquinimod.^88,89^ *CD74*, upregulated across non–T cells, supported CTCL-enriched *MIF–CD74* and *CD74–CXCR4* interactions—known STAT3 activators in other cancers and targetable with agents like STRO-001.^69,90^ Notably, *Staphylococcus aureus* superantigens may amplify STAT3 signaling, exacerbating immune evasion and drug resistance.^91^ STAT3 activation has also been associated with filaggrin downregulation in CTCL, linking it to both immune and barrier dysfunction.^92^ Prior studies have reported success with JAK inhibitors in CTCL, although with limited efficacy in MF/SS and potential relapse in immunosuppressed patients.^93^ While these findings nominate *IL-10, S100A8/A9,* and *CD74*-driven STAT3 signaling as convergent mechanisms sustaining the immunosuppressive niche in CTCL, their precise functional role in SS remains unclear, and further validation is needed to determine whether biomarker-driven or combination targeting strategies would aid therapy or compromise immune surveillance.

Despite the depth of our multimodal single-cell analysis, the modest cohort size limited detection of treatment- or outcome-specific associations, and sample pooling constrained clinical annotation. Inferred immune interactions, drawn from expression data and parallels in other cancers, require further validation to clarify their role in CTCL. Future studies incorporating longitudinal and spatial sampling will be essential to confirm malignant subtype dynamics and advance these findings toward clinical application.

In summary, we delineate three MTC subtypes in Sézary syndrome—MTC CM, MTC E/EM, and MTC Reg—each with distinct transcriptional features and therapeutic relevance. This study provides one of the most comprehensive characterizations to date of MTCs and their immunologic milieu in CTCL, laying the groundwork for deeper investigation and personalized mechanism-driven treatment approaches, and suggests a critical need to further explore how targeting both the malignant phenotype and the immune microenvironment may reshape outcomes in CTCL.

## Acknowledgements

We thank the patients and families who participated in this study and the clinical and laboratory teams involved in patient care and sample processing. We are also grateful to the investigators who generated and shared publicly available datasets that contributed to this analysis.

## Data Availability Statement

Single-cell RNA-sequencing data from UT Southwestern samples will be uploaded to GEO and made public at the time of publication. Publicly available datasets used in this study can be accessed from the corresponding publications and repositories as cited.

## Ethics Statement

Samples collected at UT Southwestern were obtained under institutional review board (IRB) approval STU 052018-005. Publicly available external datasets were obtained in compliance with ethical guidelines established by the respective institutions and approved IRBs.

