## Supplemental Figures for "Single-Cell Profiling Reveals Targetable Malignant T-Cell Subtypes and Immune Evasion Pathways in Sézary Syndrome"

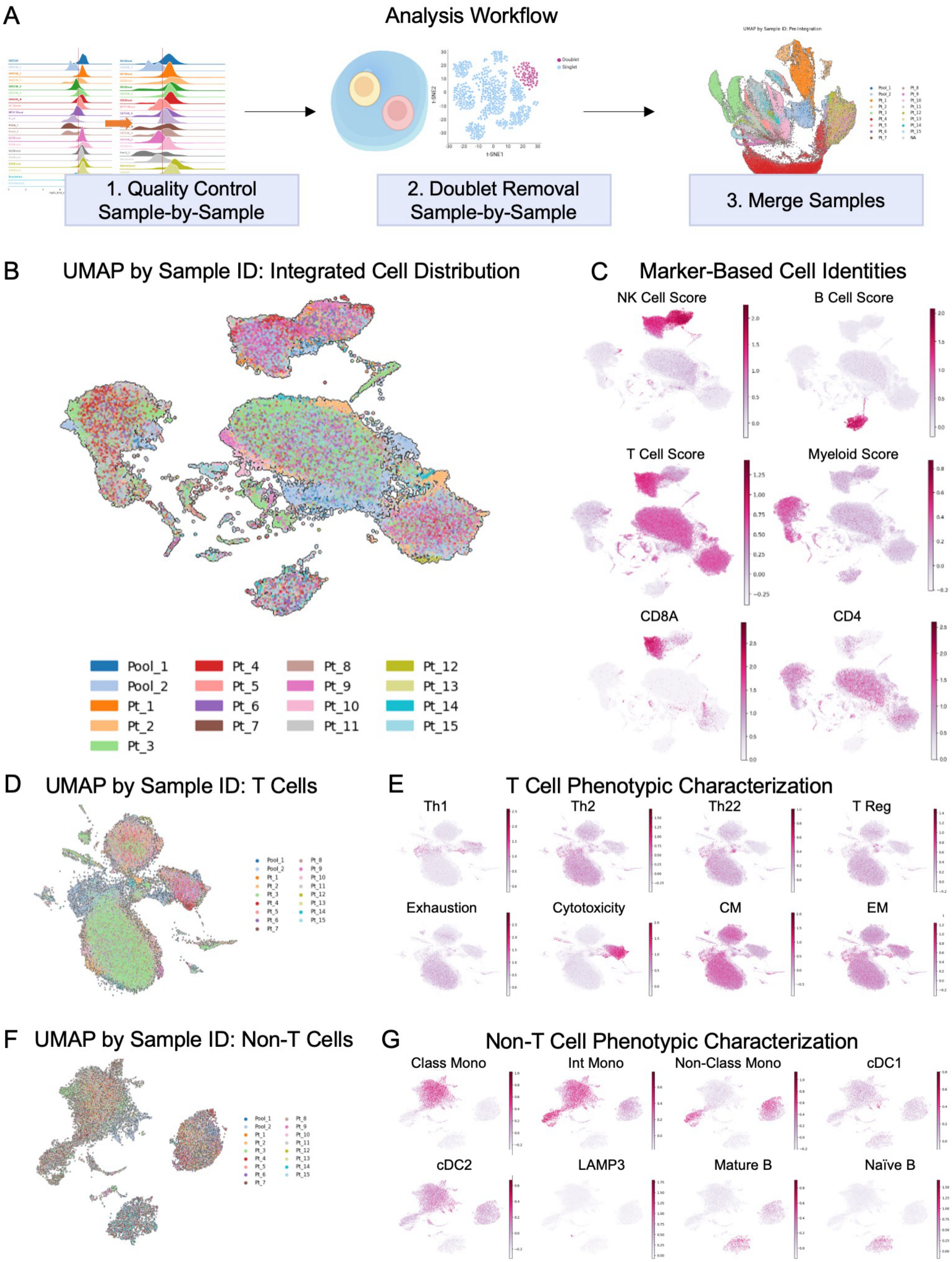
A
Analysis Workflow
1. Quality Control
Sample-by-Sample
2. Doublet Removal
Sample-by-Sample
3. Merge Samples
B
UMAP by Sample ID: Integrated Cell Distribution
Pool_1
Pool_2
Pt_1
Pt_2
Pt_3
Pt_4
Pt_5
Pt_6
Pt_7
Pt_8
Pt_9
Pt_10
Pt_11
Pt_12
Pt_13
Pt_14
Pt_15
C
Marker-Based Cell Identities
NK Cell Score
B Cell Score
T Cell Score
Myeloid Score
CD8A
CD4
D
UMAP by Sample ID: T Cells
E
T Cell Phenotypic Characterization
Th1
Th2
Th22
T Reg
Exhaustion
Cytotoxicity
CM
EM
F
UMAP by Sample ID: Non-T Cells
G
Non-T Cell Phenotypic Characterization
Class Mono
Int Mono
Non-Class Mono
cDC1
cDC2
LAMP3
Mature B
Naïve B

**Supplementary Figure 1: Quality control and data integration**

(a) Schematic representation of quality control (QC) workflow, including sample-by-sample QC via ridge plots, doublet removal, merging, and integration using Harmony. (b) UMAP embedding post-Harmony integration, colored by patient sample IDs, illustrating effective data integration. (c) UMAP plots of the entire dataset highlighting lineage-specific gene expression scores (NK, B cells, T cells, myeloid cells) and canonical markers CD8A and CD4. (d) UMAP visualization of integrated T cells colored by patient IDs, demonstrating cell distribution across samples. (e) UMAP plots displaying T-cell phenotype-specific expression scores (CM, EM, Th1, Th2, Th22, Tregs, exhaustion). (f) UMAP of integrated non-T cells colored by patient sample IDs, highlighting distribution across samples. (g) UMAP plots demonstrating phenotype-specific expression scores, identifying subsets such as classical, intermediate, non-classical monocytes, dendritic cells, and rare populations.

*Abbreviations: Th, T-helper cell; T Reg, T-regulatory cell; CM, central memory; EM, effector memory; Class Mono, classical monocyte; Int Mono, intermediate monocyte; Non-Class Mono, non-classical monocyte; cDC1, type 1 conventional dendritic cell; cDC2, type 2 conventional dendritic cell; Mature B, mature B cell; Naïve B, naïve B cell.*

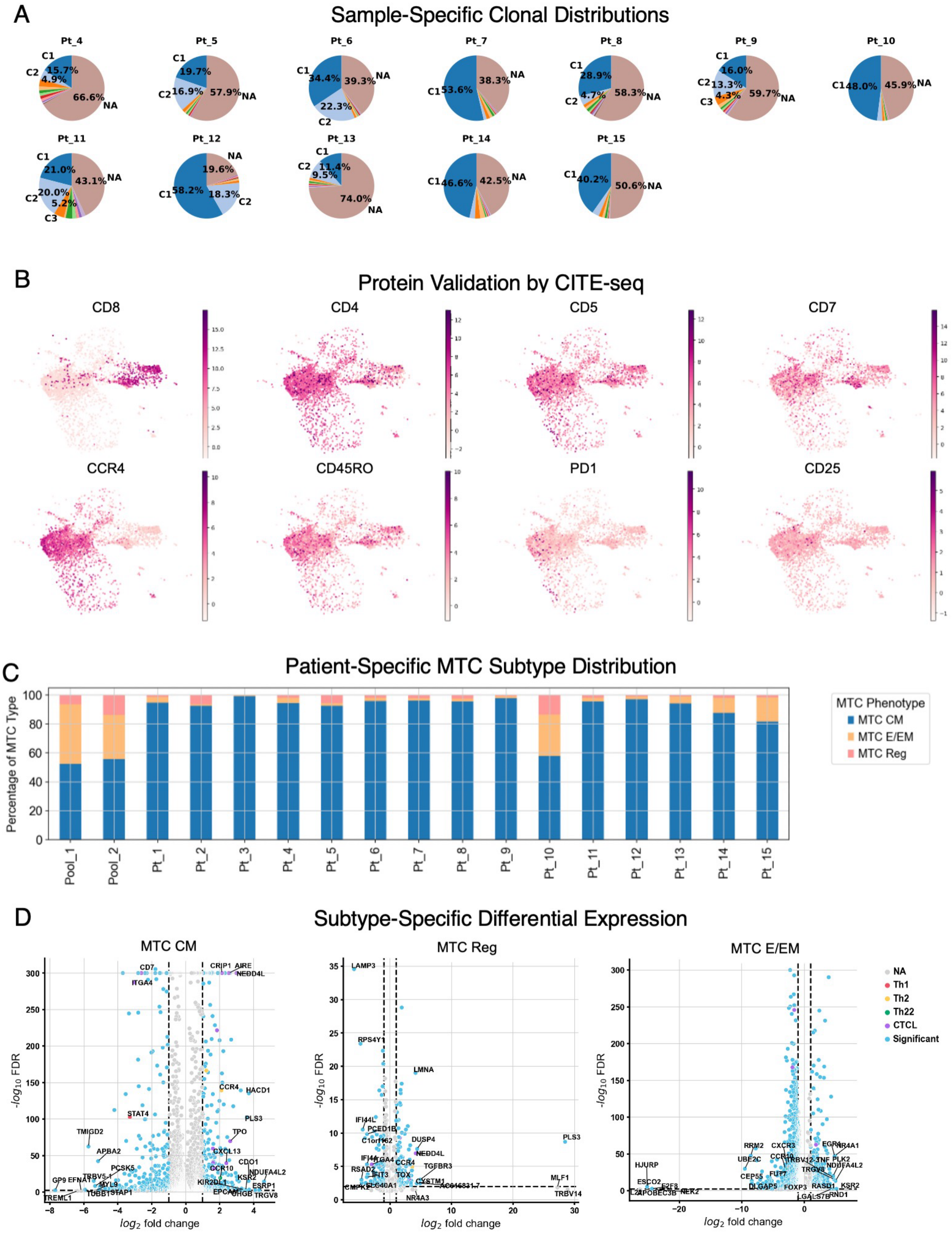


**Supplementary Figure 2: Characterization of malignant T cells**

(a) Pie charts displaying clonal distribution within individual patient samples, highlighting up to two dominant T-cell clones per patient. (b) UMAP plots from CITE-seq data validating protein expression of selected markers (CD8, CD4, CD5, CD7, CCR4, CD45RO, PD1, CD25) in comparison to scRNA-seq data (limited to UT Southwestern samples). (c) Bar graph illustrating variability in the proportion of malignant T-cell subtypes across individual patient samples. (d) Volcano plots showing differentially expressed genes in malignant T-cell subtypes versus their benign counterparts (MTC CM vs. T CM, MTC E/EM vs. T EM, MTC Reg vs. T Reg).

*Abbreviations: Th, T-helper cell; T Reg, T-regulatory cell; CM, central memory; EM, effector memory; MTC, malignant T cell; MTC CM, malignant central memory T cell; MTC E/EM, malignant effector/effector memory T cell; MTC Reg, malignant regulatory T cell; UMAP, Uniform Manifold Approximation and Projection; CITE-seq, Cellular Indexing of Transcriptomes and Epitopes by Sequencing; scRNA-seq, single-cell RNA sequencing.*

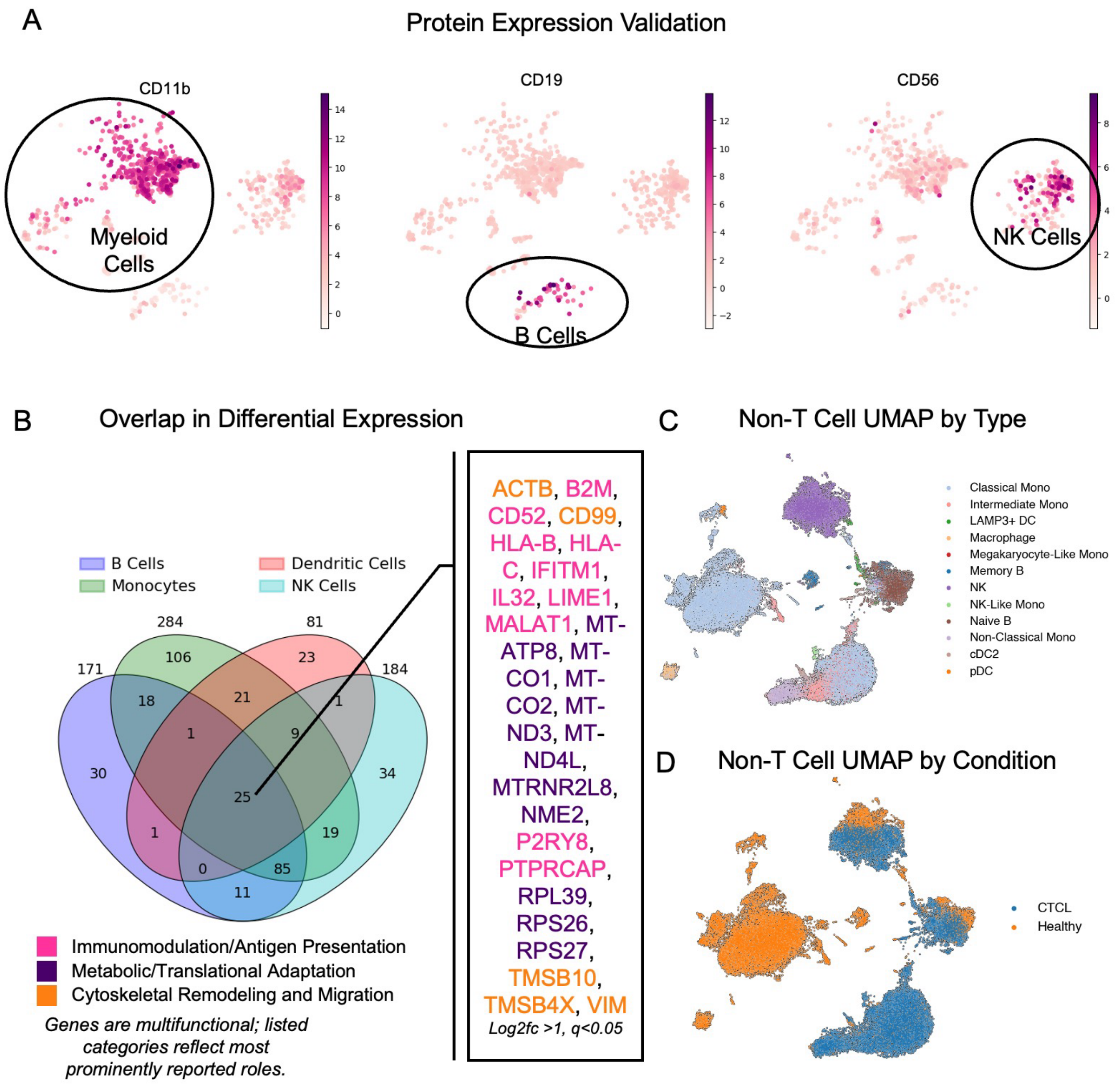


**Supplementary Figure 3: Comparative analysis of non-T cells between CTCL and healthy controls**

(a) UMAP feature plots of CITE-seq-derived surface proteins CD11b, CD19, and CD56 projected onto transcriptomic UMAP, validating non-T cell subset definitions. (b) Venn diagram summarizing overlaps in differentially expressed genes across B cells, DCs, monocytes, and NK cells, with highlighted genes significantly upregulated in both groups.

(c) UMAP visualization distinguishing non-T cell subsets across healthy control and CTCL samples. (d) UMAP visualization showing clustering of non-T cell populations by disease condition, highlighting distinct clustering of classical monocytes in CTCL.

*Abbreviations: UMAP, Uniform Manifold Approximation and Projection; CITE-seq, Cellular Indexing of Transcriptomes and Epitopes by Sequencing; CTCL, cutaneous T-cell lymphoma; NK, natural killer cells; DC, dendritic cells; Mono, monocytes; cDC2, type 2 conventional dendritic cells; DCs, dendritic cells; Memory B, memory B cells; Naïve B, naïve B cells.*

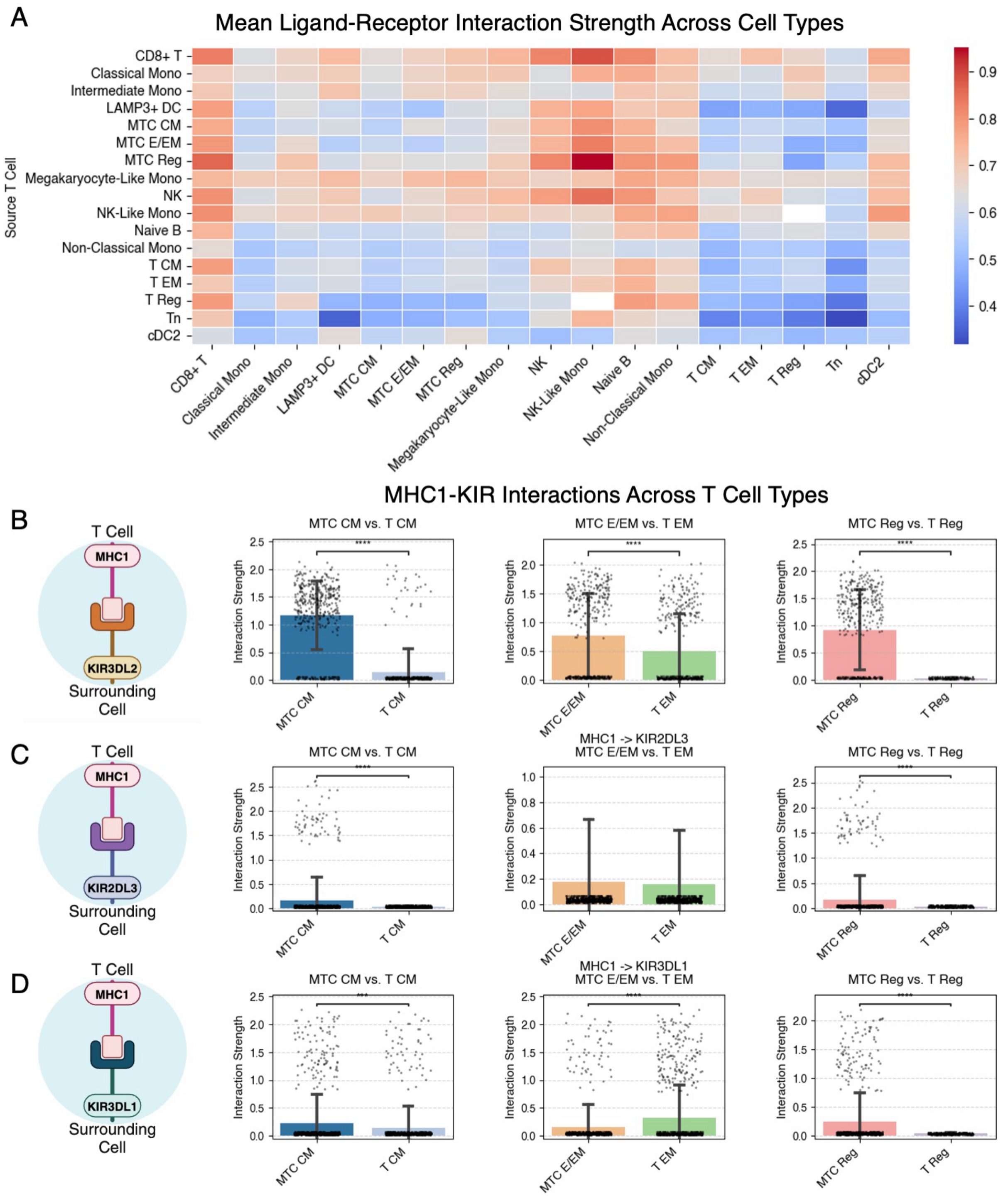


**Supplementary Figure 4: Expanded analysis of interactions and MHC I-KIR activity in MTCs**

(a) Heatmap of ligand-receptor interaction strength (lr_means), highlighting malignant T cells as primary sources of enhanced MHC class I interactions with KIR-family receptors. (b) Boxplots confirming significantly stronger KIR3DL2 interactions across all malignant subtypes (MTC CM, MTC E/EM, MTC Reg), emphasizing its relevance in immune evasion and therapeutic potential. *Statistical significance is annotated as: * = $p \leq 0.05$, ** = $p \leq 0.01$, *** = $p \leq 0.001$, **** = $p \leq 0.0001$.*

(c) Boxplots comparing interaction strengths between malignant and benign T-cell subsets, demonstrating significant elevation in MTC CM and MTC Reg. (d) MHC Class I–KIR3DL1 Interactions: Boxplots highlighting significantly increased interactions in MTC CM and MTC Reg versus benign counterparts, noting elevated signaling in benign T EM compared to MTC E/EM.

*Abbreviations: KIR, Killer Cell Immunoglobulin Like Receptor; MTC, malignant T cell; MTC CM, malignant central memory T cell; T CM, benign central memory T cell; MTC E/EM, malignant effector/effector memory T cel; T EM, benign effector memory T cell; MTC Reg, malignant regulatory T cell; T Reg, benign T-regulatory cell; MHC, major histocompatibility complex; KIR, killer immunoglobulin-like receptor.*

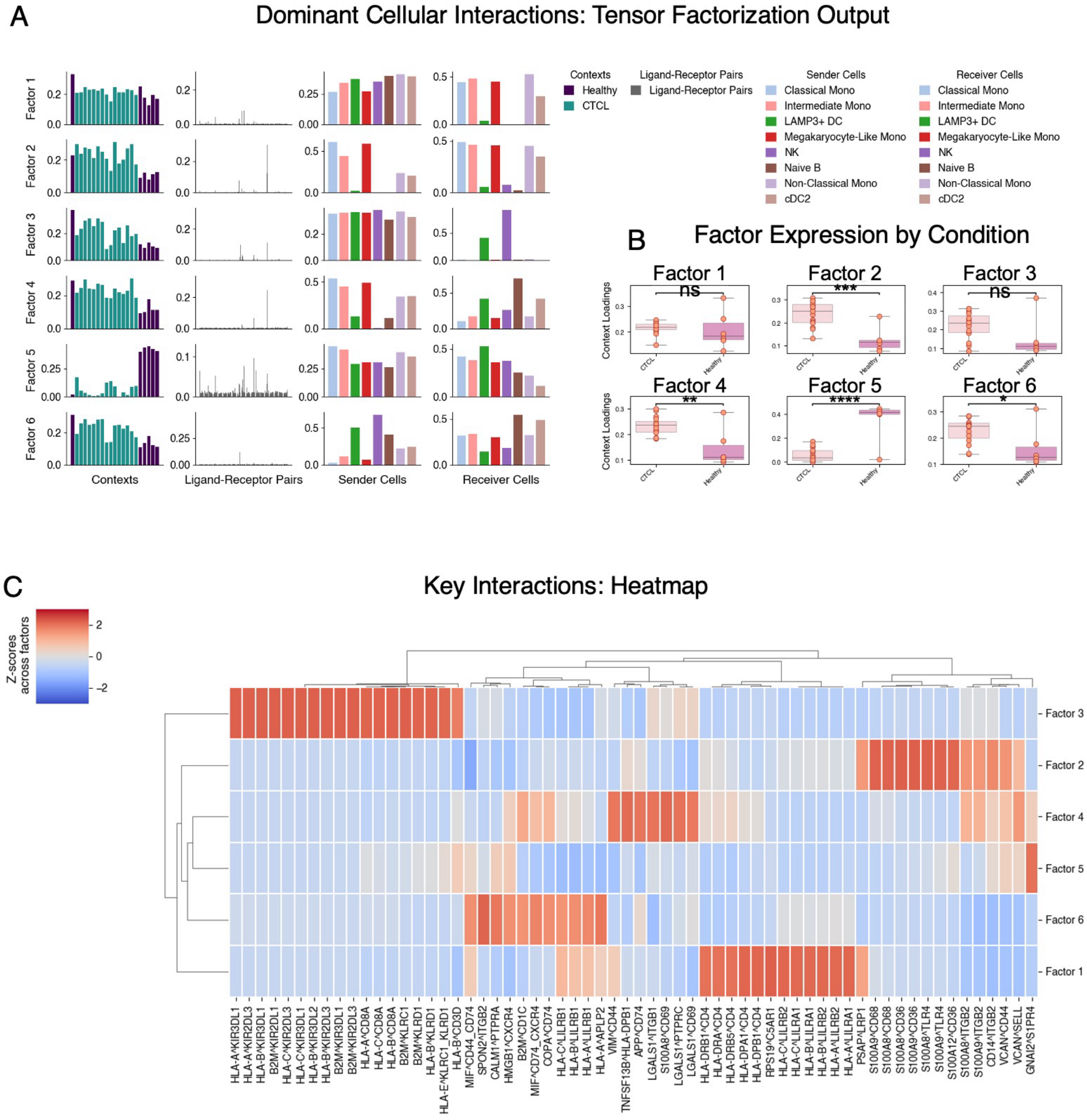


#### Supplementary Figure 5: Tensor Factorization Identifies Dominant Signaling Patterns

(a) Tensor factorization results (LIANA) illustrating major communication themes across cell types. (b) Boxplots showing differences in tensor factor expression between CTCL patients and healthy controls. Statistical significance is annotated as: ns = nonsignificant at $\alpha < 0.05$, * = $p \leq 0.05$, ** = $p \leq 0.01$, *** = $p \leq 0.001$, **** = $p \leq 0.0001$. (c) Heatmap detailing the strongest ligand-receptor interactions identified within each tensor factor.

*Abbreviations: CTCL, cutaneous T-cell lymphoma; NK, natural killer cells; DC, dendritic cells; Mono, monocytes; cDC2, type 2 conventional dendritic cells; DCs, dendritic cells; cDC2, type 2 conventional dendritic cells; Naïve B, naïve B cells.*

### SUPPLEMENTARY TABLES

| Cluster | HC_1 | HC_2 | HC_3 | HC_4 | HC_5 | HC_6 | HC_7 | Pool_1 | Pool_2 | Pt_1 | Pt_2 | Pt_3 | Pt_4 | Pt_5 | Pt_6 | Pt_7 | Pt_8 | Pt_9 | Pt_10 | Pt_11 | Pt_12 | Pt_13 | Pt_14 | Pt_15 |
|---|---|---|---|---|---|---|---|---|---|---|---|---|---|---|---|---|---|---|---|---|---|---|---|---|
| **CD8+ T** | 296 | 509 | 1093 | 0 | 221 | 1 | 396 | 130 | 457 | 337 | 171 | 388 | 715 | 35 | 131 | 129 | 115 | 1071 | 243 | 256 | 55 | 346 | 156 | 318 |
| **Class Mono** | 1348 | 6445 | 3122 | 252 | 1899 | 1384 | 872 | 4 | 255 | 312 | 16 | 1132 | 453 | 132 | 180 | 692 | 1207 | 5 | 52 | 1951 | 294 | 22 | 86 | 106 |
| **Int Mono** | 149 | 212 | 34 | 0 | 22 | 3 | 36 | 34 | 361 | 32 | 0 | 124 | 93 | 29 | 25 | 120 | 208 | 3 | 4 | 614 | 110 | 3 | 53 | 41 |
| **LAMP3+ DC** | 0 | 0 | 0 | 0 | 0 | 0 | 0 | 6 | 24 | 1 | 17 | 61 | 3 | 0 | 1 | 1 | 5 | 55 | 15 | 2 | 1 | 3 | 8 | 15 |
| **MTC CM** | 0 | 0 | 0 | 0 | 0 | 0 | 0 | 152 | 1581 | 72 | 2804 | 20377 | 276 | 101 | 411 | 486 | 645 | 1348 | 887 | 293 | 803 | 298 | 672 | 866 |
| **MTC E/EM** | 0 | 0 | 0 | 0 | 0 | 0 | 0 | 120 | 868 | 3 | 30 | 139 | 11 | 2 | 9 | 8 | 13 | 19 | 439 | 7 | 15 | 15 | 78 | 177 |
| **MTC Reg** | 0 | 0 | 0 | 0 | 0 | 0 | 0 | 18 | 386 | 1 | 192 | 38 | 5 | 6 | 8 | 11 | 16 | 12 | 206 | 6 | 9 | 3 | 15 | 17 |
| **MP** | 3 | 391 | 7 | 0 | 22 | 14 | 0 | 1 | 1 | 0 | 0 | 0 | 0 | 0 | 0 | 0 | 0 | 0 | 0 | 1 | 0 | 0 | 0 | 0 |
| **ML Mono** | 0 | 0 | 0 | 0 | 0 | 0 | 0 | 0 | 0 | 2 | 0 | 43 | 16 | 25 | 13 | 102 | 6 | 0 | 1 | 94 | 13 | 1 | 0 | 5 |
| **Memory B** | 130 | 89 | 171 | 0 | 213 | 2 | 118 | 0 | 0 | 0 | 0 | 0 | 0 | 0 | 0 | 0 | 0 | 0 | 0 | 0 | 0 | 0 | 0 | 0 |
| **NK** | 192 | 462 | 1951 | 0 | 340 | 18 | 290 | 91 | 201 | 611 | 51 | 223 | 284 | 56 | 150 | 361 | 70 | 702 | 241 | 383 | 41 | 418 | 340 | 97 |
| **NK-Like Mono** | 0 | 0 | 0 | 0 | 0 | 0 | 0 | 0 | 0 | 3 | 0 | 22 | 2 | 0 | 1 | 8 | 0 | 0 | 0 | 75 | 0 | 0 | 0 | 0 |
| **Naive B** | 258 | 186 | 390 | 0 | 439 | 0 | 165 | 18 | 32 | 2 | 13 | 39 | 174 | 58 | 150 | 75 | 0 | 80 | 230 | 24 | 44 | 229 | 513 | 120 |
| **NC Mono** | 0 | 0 | 0 | 0 | 0 | 0 | 0 | 1 | 65 | 12 | 3 | 183 | 86 | 54 | 29 | 316 | 108 | 23 | 17 | 555 | 53 | 48 | 9 | 52 |
| **T CM** | 776 | 435 | 249 | 0 | 101 | 2 | 1275 | 11 | 199 | 59 | 3290 | 2378 | 531 | 106 | 161 | 199 | 106 | 1064 | 852 | 84 | 134 | 385 | 184 | 563 |
| **T EM** | 150 | 160 | 49 | 0 | 37 | 0 | 586 | 10 | 79 | 29 | 475 | 535 | 22 | 2 | 7 | 54 | 173 | 28 | 21 | 65 | 45 | 19 | 267 | 72 |
| **T Reg** | 194 | 95 | 135 | 0 | 32 | 8 | 194 | 2 | 6 | 0 | 24 | 1 | 41 | 0 | 1 | 2 | 1 | 13 | 5 | 0 | 0 | 4 | 8 | 4 |
| **Tn** | 43 | 124 | 298 | 0 | 49 | 0 | 39 | 8 | 47 | 1 | 174 | 65 | 27 | 2 | 8 | 10 | 9 | 48 | 137 | 4 | 52 | 23 | 35 | 70 |
| **cDC2** | 0 | 1 | 0 | 0 | 0 | 0 | 76 | 4 | 29 | 6 | 2 | 35 | 19 | 10 | 25 | 26 | 12 | 12 | 3 | 65 | 7 | 24 | 7 | 24 |
| **pDC** | 7 | 21 | 43 | 0 | 27 | 2 | 10 | 0 | 0 | 0 | 0 | 0 | 0 | 0 | 0 | 0 | 0 | 0 | 0 | 0 | 0 | 0 | 0 | 0 |

#### Table S1. Cell counts per cluster and sample after quality control (QC) filtering

*CD8+ T: cytotoxic CD8+ T cells, Class Mono: classical monocytes, Int Mono: intermediate monocytes, DC: dendritic cells, MTC: malignant T cells, CM: central memory, E/EM: effector/memory, Reg: regulatory, MP: macrophage, ML Mono: megakaryocyte-like monocytes, B: B cells, NK: natural killer cells, NK-Like Mono: natura killer cell-like monocytes, NC Mono: nonclassical monocytes, Tn: T-naïve cells, cDC2: type 2 conventional dendritic cells, pDC plasmacytoid dendritic cells, cDC2 type 2 conventional dendritic cells*

| **Score Categories** | ***Marker Genes*** |
|---|---|
| NK Cell | *NKG7, GNLY, KLRD1, NCAM1, CD244* |
| Myeloid Cell | *CD68, ITGAM, CD24, ADGRE1, ITGAE* |
| B Cell | *MS4A1, CD19, CD79A, CD79B* |
| T Cell | *CD3D, CD3E, CD3G, CD4, CD8A, CD7, CD5* |
| Th1 | *GADD45B, IFNG, STAT4, TNF* |
| Th2 | *CCR6, GATA3* |
| Th22 | *AHR, CCR4, CCR6, CCR10, IL22* |
| T Reg | *FOXP3, IL12RA, CTLA4, TIGIT* |
| T Cell Exhaustion | *CXCL13, TIGIT* |
| T Cell Cytotoxicity | *GZMA, PRF1, CCL3, CCL4, IFNG, GZMB, NKG7, GZMB, GNLY* |
| T Central Memory | *CCR7, SELL, TCF7, PTPRC* |
| T Effector Memory | *CCR4, CD69, ITGAE, PTPRC, FAS* |
| Classical Monocyte | *CCR2, CD14, CCR5* |
| Intermediate Monocyte | *CD68, CCR2, CD14, CCR5, FCGR3A, FCGR3B* |
| Non-Classical Monocyte | *FCGR3A, FCGR3B, CXCR1, CX3CR1* |
| cDC1 | *BTLA, CADM1, ITGAE, ITGAX* |
| cDC2 | *CD14, CD163, CLEC10A, NOTCH2, CX3CR1, CD1C, CD2, ID2, IRF4, KLF4* |
| Mature B Cell | *CD38, CR2, FCER2* |
| Naïve B Cell | *IGHD, CD19* |

**Table S2. Gene sets used for cell type and functional state scoring (related to Supplementary Figure 1C, 1E, and 1G)**

Table S2 lists the gene sets used to calculate cell type–specific and functional state scores shown in Supplementary Figure 1C, 1E, and 1G. Scores were computed by averaging expression of marker genes previously validated in the literature for each category. *Acronyms: NK, natural killer; Th, T helper; Treg, regulatory T celll; DC, dendritic cell; cDC1/cDC2, conventional dendritic cell type 1/2.*

| Cluster | Gene | p_val | avg_log2FC | p_val_adj |
|---|---|---|---|---|
| CD8+ T | CCL5 | 0 | 4.742620468 | 0 |
| CD8+ T | NKG7 | 0 | 4.555574894 | 0 |
| CD8+ T | GZMA | 0 | 3.732661724 | 0 |
| CD8+ T | CST7 | 0 | 2.960759878 | 0 |
| CD8+ T | GZMH | 0 | 4.707960129 | 0 |
| CD8+ T | CTSW | 0 | 3.122534275 | 0 |
| CD8+ T | PRF1 | 0 | 3.268373489 | 0 |
| CD8+ T | FGFBP2 | 0 | 3.905147076 | 0 |
| CD8+ T | GZMB | 0 | 3.408941984 | 0 |
| CD8+ T | HCST | 0 | 1.606443405 | 0 |
| CD8+ T | GZMM | 0 | 2.790809631 | 0 |
| CD8+ T | GNLY | 0 | 3.311241865 | 0 |
| CD8+ T | CD8A | 0 | 4.790156841 | 0 |
| CD8+ T | CCL4 | 0 | 3.226287365 | 0 |
| CD8+ T | KLRD1 | 0 | 3.234349251 | 0 |
| CD8+ T | CD2 | 0 | 2.02773428 | 0 |
| CD8+ T | HOPX | 0 | 2.516045332 | 0 |
| CD8+ T | CD8B | 0 | 4.306833744 | 0 |
| CD8+ T | C12orf75 | 0 | 2.066260099 | 0 |
| CD8+ T | KLRG1 | 0 | 3.430784225 | 0 |
| CD8+ T | RARRES3 | 0 | 1.571729064 | 0 |
| CD8+ T | LYAR | 0 | 2.305831909 | 0 |
| CD8+ T | SYNE2 | 0 | 1.563319564 | 0 |
| CD8+ T | SYNE1 | 0 | 2.027023554 | 0 |
| CD8+ T | UBB | 0 | 0.984384835 | 0 |
| Class Mono | S100A8 | 0 | 5.319198132 | 0 |
| Class Mono | S100A9 | 0 | 4.986746311 | 0 |
| Class Mono | S100A12 | 0 | 5.884672642 | 0 |
| Class Mono | LYZ | 0 | 5.227620602 | 0 |
| Class Mono | MNDA | 0 | 5.333661556 | 0 |
| Class Mono | FCN1 | 0 | 5.050788879 | 0 |
| Class Mono | CTSS | 0 | 3.418771029 | 0 |
| Class Mono | VCAN | 0 | 5.776087761 | 0 |
| Class Mono | LST1 | 0 | 4.009200096 | 0 |
| Class Mono | MS4A6A | 0 | 5.210998058 | 0 |
| Class Mono | CSTA | 0 | 5.161288261 | 0 |
| Class Mono | AIF1 | 0 | 4.098974228 | 0 |
| Class Mono | GRN | 0 | 3.655825138 | 0 |

| | | | | |
|---|---|---|---|---|
| Class Mono | SERPINA1 | 0 | 4.160812378 | 0 |
| Class Mono | CST3 | 0 | 3.873757362 | 0 |
| Class Mono | TYMP | 0 | 3.534389734 | 0 |
| Class Mono | SPI1 | 0 | 4.144917488 | 0 |
| Class Mono | NCF2 | 0 | 4.928340912 | 0 |
| Class Mono | TKT | 0 | 2.821470499 | 0 |
| Class Mono | TALDO1 | 0 | 2.789353371 | 0 |
| Class Mono | CD14 | 0 | 5.298225403 | 0 |
| Class Mono | TYROBP | 0 | 3.517752647 | 0 |
| Class Mono | PSAP | 0 | 2.625902176 | 0 |
| Class Mono | CEBPD | 0 | 3.806642294 | 0 |
| Class Mono | APLP2 | 0 | 3.678227186 | 0 |
| Int Mono | CST3 | 0 | 3.925885439 | 0 |
| Int Mono | SAT1 | 0 | 2.802797794 | 0 |
| Int Mono | HLA-DRA | 0 | 3.576267958 | 0 |
| Int Mono | PSAP | 0 | 2.739881516 | 0 |
| Int Mono | TYROBP | 0 | 3.314064503 | 0 |
| Int Mono | AIF1 | 0 | 3.19081378 | 0 |
| Int Mono | FCER1G | 0 | 2.936057806 | 0 |
| Int Mono | HLA-DRB1 | 0 | 3.432316065 | 0 |
| Int Mono | FCN1 | 0 | 3.259858608 | 0 |
| Int Mono | LYZ | 0 | 3.276810408 | 0 |
| Int Mono | SERPINA1 | 0 | 3.029562235 | 0 |
| Int Mono | CTSS | 0 | 2.593741894 | 0 |
| Int Mono | SPI1 | 0 | 2.725101948 | 0 |
| Int Mono | LST1 | 0 | 2.776778221 | 0 |
| Int Mono | NEAT1 | 0 | 2.320338726 | 0 |
| Int Mono | TYMP | 0 | 2.538033962 | 0 |
| Int Mono | CD74 | 0 | 3.025821209 | 0 |
| Int Mono | HLA-DPA1 | 0 | 2.850233793 | 0 |
| Int Mono | S100A8 | 0 | 3.552746058 | 0 |
| Int Mono | FTL | 0 | 2.686908007 | 0 |
| Int Mono | CFD | 0 | 2.870903492 | 0 |
| Int Mono | FTH1 | 0 | 2.44031477 | 0 |
| Int Mono | S100A9 | 0 | 3.122757912 | 0 |
| Int Mono | HLA-DPB1 | 0 | 2.721873999 | 0 |
| Int Mono | NPC2 | 0 | 2.133535385 | 0 |
| LAMP3+ DC | MALAT1 | 1.32E-22 | 1.27797246 | 2.971E-18 |
| LAMP3+ DC | IL32 | 3.92E-22 | 1.396389842 | 4.396E-18 |
| LAMP3+ DC | RPS26 | 1.07E-16 | 0.794490278 | 5.975E-13 |
| LAMP3+ DC | MTRNR2L12 | 3.84E-15 | 1.907307863 | 1.229E-11 |

| | | | | |
|---|---|---|---|---|
| LAMP3+ DC | B2M | 8.25E-15 | 0.891773283 | 2.124E-11 |
| LAMP3+ DC | TMSB10 | 1.12E-14 | 0.771938741 | 2.512E-11 |
| LAMP3+ DC | CD52 | 1.78E-14 | 0.914542675 | 3.621E-11 |
| LAMP3+ DC | VIM | 1.29E-13 | 0.679937482 | 1.7E-10 |
| LAMP3+ DC | CD3E | 2.68E-13 | 0.921808064 | 2.868E-10 |
| LAMP3+ DC | RPS21 | 4.84E-13 | 0.516103804 | 4.522E-10 |
| LAMP3+ DC | RPL41 | 4.02E-12 | 0.672854125 | 2.917E-09 |
| LAMP3+ DC | RPS12 | 4.14E-12 | 0.661908209 | 2.917E-09 |
| LAMP3+ DC | POTEE | 5.71E-12 | 7.720767021 | 3.746E-09 |
| LAMP3+ DC | IFITM1 | 8.52E-12 | 0.929033637 | 5.04E-09 |
| LAMP3+ DC | EEF1A1 | 2.15E-11 | 0.829662204 | 1.099E-08 |
| LAMP3+ DC | MT-ND4L | 3E-11 | 0.79827863 | 1.375E-08 |
| LAMP3+ DC | RPS27 | 4.21E-11 | 0.725014806 | 1.85E-08 |
| LAMP3+ DC | RPS28 | 4.47E-11 | 0.550162613 | 1.93E-08 |
| LAMP3+ DC | PTMA | 9.07E-11 | 0.418745637 | 3.562E-08 |
| LAMP3+ DC | RPL36 | 9.21E-11 | 0.477515608 | 3.562E-08 |
| LAMP3+ DC | RPS8 | 1.26E-10 | 0.58488971 | 4.799E-08 |
| LAMP3+ DC | RPS15A | 5.61E-10 | 0.563026428 | 1.879E-07 |
| LAMP3+ DC | H3F3B | 7.81E-10 | 0.391081452 | 2.502E-07 |
| LAMP3+ DC | RPS3A | 1.37E-09 | 0.494040638 | 4.206E-07 |
| LAMP3+ DC | RPS29 | 5.33E-09 | 0.441179752 | 1.358E-06 |
| MTC CM | CD52 | 0 | 2.351050615 | 0 |
| MTC CM | IL32 | 0 | 2.892216921 | 0 |
| MTC CM | S100A10 | 0 | 1.966946721 | 0 |
| MTC CM | CRIP1 | 0 | 2.835954905 | 0 |
| MTC CM | TMSB10 | 0 | 1.520302773 | 0 |
| MTC CM | IFITM1 | 0 | 2.144979239 | 0 |
| MTC CM | B2M | 0 | 1.672727704 | 0 |
| MTC CM | TRBV7-2 | 0 | 4.897675991 | 0 |
| MTC CM | CD3D | 0 | 2.129636526 | 0 |
| MTC CM | LIMD2 | 0 | 1.467060328 | 0 |
| MTC CM | LTB | 0 | 2.167132616 | 0 |
| MTC CM | TMSB4X | 0 | 1.435465932 | 0 |
| MTC CM | RPS4X | 0 | 1.313147664 | 0 |
| MTC CM | HINT1 | 0 | 1.206465602 | 0 |
| MTC CM | PTPRCAP | 0 | 2.380532503 | 0 |
| MTC CM | MT-CO2 | 0 | 1.27576685 | 0 |
| MTC CM | LAT | 0 | 2.255853176 | 0 |
| MTC CM | NME2 | 0 | 1.949710846 | 0 |
| MTC CM | RPS16 | 0 | 0.920181572 | 0 |
| MTC CM | RPL17 | 0 | 1.478845596 | 0 |

| | | | | |
|---|---|---|---|---|
| MTC CM | ACTG1 | 0 | 1.032556176 | 0 |
| MTC CM | RPS7 | 0 | 0.96927774 | 0 |
| MTC CM | CD99 | 0 | 1.658320785 | 0 |
| MTC CM | TAGLN2 | 0 | 1.190759182 | 0 |
| MTC CM | RPL18A | 0 | 1.127559543 | 0 |
| MTC E/EM | MTRNR2L12 | 0 | 3.797419786 | 0 |
| MTC E/EM | JUN | 0 | 3.159010172 | 0 |
| MTC E/EM | MALAT1 | 0 | 1.876743913 | 0 |
| MTC E/EM | BTG1 | 0 | 1.411910534 | 0 |
| MTC E/EM | JUNB | 0 | 1.717103004 | 0 |
| MTC E/EM | UBC | 0 | 1.393598676 | 0 |
| MTC E/EM | IER2 | 1.1E-281 | 1.934652567 | 2.75E-278 |
| MTC E/EM | PPP1R15A | 5.4E-274 | 2.62601757 | 1.2E-270 |
| MTC E/EM | H3F3B | 2.8E-269 | 1.076944113 | 5.69E-266 |
| MTC E/EM | MT-ATP6 | 4.2E-264 | 1.064448118 | 7.8E-261 |
| MTC E/EM | PTMA | 4.8E-249 | 0.833797872 | 7.17E-246 |
| MTC E/EM | HSP90AB1 | 8E-249 | 1.582884073 | 1.13E-245 |
| MTC E/EM | LMNA | 2.9E-247 | 3.13697052 | 3.88E-244 |
| MTC E/EM | NFKBIA | 1.4E-245 | 2.136007309 | 1.71E-242 |
| MTC E/EM | MT-CYB | 1.1E-239 | 0.937195718 | 1.3E-236 |
| MTC E/EM | HSP90AA1 | 4.3E-236 | 1.369690418 | 4.77E-233 |
| MTC E/EM | KLF6 | 1.3E-226 | 1.551624298 | 1.34E-223 |
| MTC E/EM | DDX5 | 5.4E-201 | 0.826834798 | 4.89E-198 |
| MTC E/EM | FOSB | 7.6E-198 | 3.108406305 | 6.59E-195 |
| MTC E/EM | EIF1 | 8.7E-190 | 0.683790624 | 7.27E-187 |
| MTC E/EM | GADD45B | 1.2E-180 | 2.795935869 | 8.7E-178 |
| MTC E/EM | BRD2 | 2.6E-177 | 2.16755271 | 1.84E-174 |
| MTC E/EM | MT-ND4L | 8.2E-177 | 1.052663088 | 5.57E-174 |
| MTC E/EM | NR4A2 | 1.2E-173 | 3.062298059 | 7.97E-171 |
| MTC E/EM | SRSF7 | 5.6E-170 | 1.784582257 | 3.42E-167 |
| MTC Reg | RPL23A | 3.6E-149 | 1.066788673 | 8.03E-145 |
| MTC Reg | MTRNR2L12 | 1.3E-142 | 2.830509901 | 1.5E-138 |
| MTC Reg | RPL41 | 9.8E-140 | 1.193085551 | 7.3E-136 |
| MTC Reg | LMNA | 4.4E-135 | 2.822323084 | 2.46E-131 |
| MTC Reg | VIM | 5.4E-116 | 1.058030844 | 2.44E-112 |
| MTC Reg | TRBC1 | 1.1E-112 | 1.977428436 | 4.3E-109 |
| MTC Reg | RGCC | 1.4E-112 | 2.283359051 | 4.5E-109 |
| MTC Reg | ITGB1 | 8.7E-107 | 1.341950178 | 2.43E-103 |
| MTC Reg | EIF1 | 7.8E-106 | 0.697488248 | 1.94E-102 |
| MTC Reg | H3F3B | 7.7E-104 | 0.776537478 | 1.62E-100 |
| MTC Reg | RPL19 | 1.6E-100 | 0.875975609 | 2.988E-97 |

| | | | | |
|---|---|---|---|---|
| MTC Reg | PTMA | 2.7E-100 | 0.73281008 | 4.61E-97 |
| MTC Reg | MALAT1 | 6.3E-99 | 1.298456192 | 1.011E-95 |
| MTC Reg | BTG1 | 5.88E-94 | 0.948036194 | 8.79E-91 |
| MTC Reg | TSHZ2 | 1.42E-93 | 1.918074965 | 1.984E-90 |
| MTC Reg | HLA-A | 1.38E-91 | 0.77121067 | 1.826E-88 |
| MTC Reg | RPS21 | 4.82E-88 | 0.774923742 | 6.013E-85 |
| MTC Reg | RNF19A | 3.89E-87 | 1.995276809 | 4.591E-84 |
| MTC Reg | CCR7 | 4.74E-85 | 1.570336938 | 5.312E-82 |
| MTC Reg | BIRC3 | 1.24E-84 | 1.797584891 | 1.326E-81 |
| MTC Reg | UBC | 2.27E-80 | 1.08375001 | 2.126E-77 |
| MTC Reg | DUSP4 | 2.22E-74 | 3.270547628 | 1.917E-71 |
| MTC Reg | EEF1A1 | 2.24E-72 | 0.985729277 | 1.795E-69 |
| MTC Reg | PLS3 | 8.27E-72 | 2.930764437 | 6.395E-69 |
| MTC Reg | RPL38 | 9.46E-70 | 0.570886791 | 7.078E-67 |
| MP | GPX1 | 4.2E-237 | 3.447975397 | 9.44E-233 |
| MP | CTSB | 8.8E-219 | 2.760019541 | 9.83E-215 |
| MP | MS4A6A | 9.4E-203 | 3.228405476 | 7.05E-199 |
| MP | CD163 | 5.9E-201 | 3.605788946 | 3.32E-197 |
| MP | BLVRB | 1.8E-172 | 2.970825911 | 4.41E-169 |
| MP | ZYX | 1.7E-169 | 2.308404446 | 3.71E-166 |
| MP | GABARAP | 1.6E-166 | 2.213089228 | 2.7E-163 |
| MP | HMOX1 | 4E-164 | 3.77559495 | 6.4E-161 |
| MP | LST1 | 7.9E-161 | 2.646500349 | 1.11E-157 |
| MP | SPI1 | 3.8E-160 | 2.763626337 | 5.07E-157 |
| MP | STXBP2 | 1.7E-158 | 2.69820118 | 2.08E-155 |
| MP | AIF1 | 4.2E-157 | 2.640525818 | 4.91E-154 |
| MP | PGD | 1.1E-155 | 2.821619511 | 1.2E-152 |
| MP | TYMP | 1.1E-155 | 2.450615406 | 1.2E-152 |
| MP | FGR | 3.3E-155 | 2.715561867 | 3.32E-152 |
| MP | NPC2 | 6.6E-154 | 2.17496419 | 6.43E-151 |
| MP | IFI30 | 2.7E-152 | 4.126478672 | 2.51E-149 |
| MP | PSAP | 5E-152 | 1.884925008 | 4.53E-149 |
| MP | RNF130 | 5.1E-151 | 2.830433607 | 4.4E-148 |
| MP | PYCARD | 4.7E-150 | 2.258198023 | 3.92E-147 |
| MP | VCAN | 4E-147 | 2.791413069 | 3.09E-144 |
| MP | SULT1A1 | 1.3E-146 | 3.596722603 | 9.48E-144 |
| MP | C1orf162 | 5.5E-146 | 2.302686214 | 3.97E-143 |
| MP | CST3 | 1.4E-144 | 2.043902636 | 9.61E-142 |
| MP | LILRB2 | 1.4E-144 | 3.395792961 | 9.69E-142 |
| ML Mono | LYZ | 1.6E-165 | 4.437789917 | 3.5E-161 |
| ML Mono | S100A9 | 1.3E-161 | 5.042582035 | 1.4E-157 |

| | | | | |
|---|---|---|---|---|
| ML Mono | S100A8 | 4.3E-159 | 5.525038719 | 3.19E-155 |
| ML Mono | FTL | 2.5E-157 | 3.278684378 | 1.38E-153 |
| ML Mono | TYROBP | 4.9E-156 | 3.785999298 | 2.21E-152 |
| ML Mono | CST3 | 2.2E-155 | 4.109960079 | 8.25E-152 |
| ML Mono | FTH1 | 2.2E-143 | 2.407633066 | 7.14E-140 |
| ML Mono | FCN1 | 2.8E-138 | 3.503867388 | 7.83E-135 |
| ML Mono | S100A6 | 6.9E-134 | 2.193773031 | 1.72E-130 |
| ML Mono | PSAP | 3.2E-133 | 2.696703911 | 7.09E-130 |
| ML Mono | CTSS | 1.2E-131 | 2.842621565 | 2.41E-128 |
| ML Mono | AIF1 | 2.3E-126 | 3.121240854 | 4.22E-123 |
| ML Mono | LST1 | 9.3E-122 | 2.885006428 | 1.6E-118 |
| ML Mono | TSPO | 2E-121 | 2.106232405 | 3.13E-118 |
| ML Mono | GABARAP | 2.8E-120 | 2.308771133 | 4.13E-117 |
| ML Mono | OAZ1 | 8.2E-117 | 1.504693747 | 1.14E-113 |
| ML Mono | VCAN | 8.3E-109 | 3.049759865 | 1.1E-105 |
| ML Mono | CD74 | 5.4E-108 | 2.689528704 | 6.69E-105 |
| ML Mono | SAT1 | 3.3E-107 | 2.356158733 | 3.86E-104 |
| ML Mono | GSTP1 | 4.1E-106 | 2.298722982 | 4.58E-103 |
| ML Mono | NCF1 | 2.2E-104 | 3.123024702 | 2.3E-101 |
| ML Mono | S100A4 | 6.8E-104 | 1.915719271 | 6.93E-101 |
| ML Mono | S100A12 | 7.7E-104 | 3.082556963 | 7.51E-101 |
| ML Mono | NEAT1 | 6E-103 | 2.091389894 | 5.64E-100 |
| ML Mono | SPI1 | 1.89E-96 | 2.582244396 | 1.692E-93 |
| Memory B | CD79A | 0 | 5.871738911 | 0 |
| Memory B | MS4A1 | 0 | 6.023410797 | 0 |
| Memory B | BANK1 | 0 | 6.018186569 | 0 |
| Memory B | CD79B | 0 | 4.773616314 | 0 |
| Memory B | RALGPS2 | 0 | 5.342929363 | 0 |
| Memory B | HLA-DPB1 | 0 | 2.990559101 | 0 |
| Memory B | TNFRSF13C | 2.4E-302 | 6.182079315 | 7.78E-299 |
| Memory B | HLA-DPA1 | 7.4E-283 | 2.702003956 | 1.67E-279 |
| Memory B | HLA-DRA | 3.1E-277 | 2.852632284 | 6.42E-274 |
| Memory B | LINC00926 | 5.5E-269 | 5.868813038 | 9.41E-266 |
| Memory B | IGHM | 1.4E-267 | 5.162686825 | 2.2E-264 |
| Memory B | FCMR | 1.5E-267 | 3.009572268 | 2.22E-264 |
| Memory B | IGKC | 2.9E-262 | 4.158593178 | 4.06E-259 |
| Memory B | HLA-DQB1 | 6.7E-255 | 3.774262905 | 8.78E-252 |
| Memory B | BLK | 1.4E-243 | 6.172082901 | 1.65E-240 |
| Memory B | IRF8 | 1.8E-237 | 3.750537872 | 2.04E-234 |
| Memory B | HLA-DMA | 2.5E-236 | 3.137788773 | 2.67E-233 |
| Memory B | POU2F2 | 1.3E-226 | 2.672762632 | 1.29E-223 |

| | | | | |
|---|---|---|---|---|
| Memory B | CD22 | 6.1E-221 | 6.092372894 | 5.94E-218 |
| Memory B | HLA-DRB1 | 3E-218 | 2.623134851 | 2.79E-215 |
| Memory B | HLA-DMB | 2.5E-216 | 3.579456329 | 2.25E-213 |
| Memory B | SWAP70 | 6.5E-206 | 4.034497261 | 5.02E-203 |
| Memory B | BCL11A | 2.9E-202 | 5.168663025 | 2.13E-199 |
| Memory B | HLA-DQA1 | 1E-201 | 3.910999537 | 7.19E-199 |
| Memory B | MEF2C | 3.4E-196 | 3.179704666 | 2.21E-193 |
| NK | NKG7 | 0 | 5.04956913 | 0 |
| NK | PRF1 | 0 | 4.82682991 | 0 |
| NK | GZMB | 0 | 5.337763309 | 0 |
| NK | CTSW | 0 | 4.419467449 | 0 |
| NK | GZMA | 0 | 4.496548176 | 0 |
| NK | KLRD1 | 0 | 4.898091793 | 0 |
| NK | GNLY | 0 | 5.039386749 | 0 |
| NK | CST7 | 0 | 3.344718695 | 0 |
| NK | HOPX | 0 | 4.2488451 | 0 |
| NK | FGFBP2 | 0 | 4.879288673 | 0 |
| NK | CD7 | 0 | 3.274960041 | 0 |
| NK | CCL5 | 0 | 3.960488558 | 0 |
| NK | KLRF1 | 0 | 5.63166666 | 0 |
| NK | CD247 | 0 | 2.65647459 | 0 |
| NK | FCGR3A | 0 | 3.573628187 | 0 |
| NK | GZMM | 0 | 3.331202745 | 0 |
| NK | CLIC3 | 0 | 5.074412346 | 0 |
| NK | SPON2 | 0 | 4.693784237 | 0 |
| NK | KLRB1 | 0 | 3.660100222 | 0 |
| NK | HCST | 0 | 1.836464763 | 0 |
| NK | CCL4 | 0 | 3.719687939 | 0 |
| NK | IL2RB | 0 | 3.737388134 | 0 |
| NK | PLAC8 | 0 | 2.48375988 | 0 |
| NK | EFHD2 | 0 | 2.105691671 | 0 |
| NK | MATK | 0 | 4.152040005 | 0 |
| NK-Like Mono | S100A8 | 3.62E-57 | 5.276485443 | 8.121E-53 |
| NK-Like Mono | S100A9 | 6.07E-51 | 3.991076231 | 6.81E-47 |
| NK-Like Mono | FTL | 2.26E-48 | 2.70349431 | 1.688E-44 |
| NK-Like Mono | TYROBP | 3.31E-46 | 3.204213381 | 1.856E-42 |
| NK-Like Mono | CST3 | 8.59E-46 | 3.368080616 | 3.855E-42 |
| NK-Like Mono | LYZ | 1.12E-45 | 3.335815191 | 4.206E-42 |
| NK-Like Mono | FTH1 | 1.99E-45 | 2.116714239 | 6.393E-42 |
| NK-Like Mono | NEAT1 | 3.31E-41 | 2.069669008 | 9.293E-38 |
| NK-Like Mono | S100A6 | 5.23E-41 | 1.988019228 | 1.303E-37 |

| | | | | |
|---|---|---|---|---|
| NK-Like Mono | FOS | 1.05E-35 | 2.520178795 | 2.366E-32 |
| NK-Like Mono | S100A4 | 2.47E-35 | 1.844208121 | 5.039E-32 |
| NK-Like Mono | GABARAP | 2.84E-35 | 2.025396585 | 5.311E-32 |
| NK-Like Mono | NKG7 | 3.4E-34 | 3.237766266 | 5.862E-31 |
| NK-Like Mono | S100A12 | 1.28E-33 | 2.78222394 | 2.051E-30 |
| NK-Like Mono | GSTP1 | 1.09E-32 | 2.125247717 | 1.634E-29 |
| NK-Like Mono | CCL5 | 4.08E-32 | 3.155453682 | 5.716E-29 |
| NK-Like Mono | FCN1 | 3.17E-31 | 2.59732008 | 4.178E-28 |
| NK-Like Mono | CD74 | 3.41E-31 | 1.945868254 | 4.248E-28 |
| NK-Like Mono | VCAN | 7.81E-30 | 2.521843195 | 9.225E-27 |
| NK-Like Mono | AIF1 | 4.82E-29 | 2.328007698 | 5.407E-26 |
| NK-Like Mono | GNLY | 9.42E-29 | 3.510245085 | 1.007E-25 |
| NK-Like Mono | MT-ND4 | 1.09E-27 | 1.176753879 | 1.112E-24 |
| NK-Like Mono | NCF1 | 2.26E-27 | 2.342805624 | 2.201E-24 |
| NK-Like Mono | TSPO | 8.63E-26 | 1.510815024 | 8.069E-23 |
| NK-Like Mono | MTRNR2L8 | 1.28E-25 | 3.490778923 | 1.147E-22 |
| Naive B | CD79A | 0 | 7.416737556 | 0 |
| Naive B | MS4A1 | 0 | 7.727255821 | 0 |
| Naive B | HLA-DPB1 | 0 | 3.800414562 | 0 |
| Naive B | HLA-DRA | 0 | 4.030269146 | 0 |
| Naive B | HLA-DPA1 | 0 | 3.649439812 | 0 |
| Naive B | CD79B | 0 | 4.754781723 | 0 |
| Naive B | CD37 | 0 | 2.156935692 | 0 |
| Naive B | BANK1 | 0 | 6.415452957 | 0 |
| Naive B | IGHM | 0 | 5.95330286 | 0 |
| Naive B | HLA-DQB1 | 0 | 4.348500729 | 0 |
| Naive B | LINC00926 | 0 | 7.004324913 | 0 |
| Naive B | HLA-DQA1 | 0 | 4.978325844 | 0 |
| Naive B | HLA-DRB1 | 0 | 3.46587348 | 0 |
| Naive B | CD74 | 0 | 3.659224033 | 0 |
| Naive B | RALGPS2 | 0 | 5.222727776 | 0 |
| Naive B | IGHD | 0 | 7.307136536 | 0 |
| Naive B | MEF2C | 0 | 3.433150768 | 0 |
| Naive B | HLA-DMA | 0 | 3.02235961 | 0 |
| Naive B | FCMR | 0 | 2.67693615 | 0 |
| Naive B | CXCR4 | 0 | 2.157906771 | 0 |
| Naive B | FAM129C | 0 | 6.644666195 | 0 |
| Naive B | FCER2 | 0 | 6.956933022 | 0 |
| Naive B | CD22 | 0 | 6.540520191 | 0 |
| Naive B | IRF8 | 0 | 3.325276136 | 0 |
| Naive B | BLK | 0 | 6.129780293 | 0 |

| | | | | |
|---|---|---|---|---|
| NC Mono | LST1 | 0 | 4.560498238 | 0 |
| NC Mono | PSAP | 0 | 3.772663116 | 0 |
| NC Mono | AIF1 | 0 | 4.217755318 | 0 |
| NC Mono | IFITM3 | 0 | 4.602684975 | 0 |
| NC Mono | FCER1G | 0 | 4.019050598 | 0 |
| NC Mono | FTH1 | 0 | 3.269052982 | 0 |
| NC Mono | SERPINA1 | 0 | 4.255123615 | 0 |
| NC Mono | SAT1 | 0 | 3.396457195 | 0 |
| NC Mono | FCGR3A | 0 | 4.565264225 | 0 |
| NC Mono | FTL | 0 | 3.651969433 | 0 |
| NC Mono | CST3 | 0 | 4.525194168 | 0 |
| NC Mono | HLA-DPA1 | 0 | 3.925172091 | 0 |
| NC Mono | TYROBP | 0 | 3.863112211 | 0 |
| NC Mono | MS4A7 | 0 | 4.153521061 | 0 |
| NC Mono | CD68 | 0 | 4.051233292 | 0 |
| NC Mono | CTSS | 0 | 3.138445616 | 0 |
| NC Mono | CD74 | 0 | 3.626972198 | 0 |
| NC Mono | S100A11 | 0 | 2.319823265 | 0 |
| NC Mono | SPI1 | 0 | 3.31670022 | 0 |
| NC Mono | HLA-DRB1 | 0 | 3.695339441 | 0 |
| NC Mono | CFD | 0 | 3.742064476 | 0 |
| NC Mono | COTL1 | 0 | 2.517810583 | 0 |
| NC Mono | HLA-DRA | 0 | 3.67037034 | 0 |
| NC Mono | OAZ1 | 0 | 1.706267595 | 0 |
| NC Mono | IFITM2 | 0 | 2.33959341 | 0 |
| T CM | RPS12 | 0 | 1.523334026 | 0 |
| T CM | RPL13 | 0 | 1.322485685 | 0 |
| T CM | RPL34 | 0 | 1.242499948 | 0 |
| T CM | EEF1A1 | 0 | 1.362612009 | 0 |
| T CM | RPL32 | 0 | 1.262604237 | 0 |
| T CM | RPS6 | 0 | 1.282004833 | 0 |
| T CM | RPS27 | 0 | 1.43744266 | 0 |
| T CM | RPS27A | 0 | 1.205642939 | 0 |
| T CM | RPS3A | 0 | 1.23378861 | 0 |
| T CM | RPS25 | 0 | 1.222782731 | 0 |
| T CM | RPS29 | 0 | 1.435167789 | 0 |
| T CM | RPL3 | 0 | 1.277033925 | 0 |
| T CM | RPS8 | 0 | 1.227128506 | 0 |
| T CM | RPL35A | 0 | 1.103004336 | 0 |
| T CM | RPS3 | 0 | 1.193499327 | 0 |
| T CM | RPL37 | 0 | 1.113551021 | 0 |

| | | | | |
|---|---|---|---|---|
| T CM | RPL39 | 0 | 1.183073163 | 0 |
| T CM | RPL10A | 0 | 1.16237855 | 0 |
| T CM | RPS18 | 0 | 1.292509198 | 0 |
| T CM | RPL9 | 0 | 1.131461859 | 0 |
| T CM | RPL30 | 0 | 1.176509142 | 0 |
| T CM | RPL36 | 0 | 1.123114586 | 0 |
| T CM | RPS23 | 0 | 1.158531547 | 0 |
| T CM | RPL5 | 0 | 1.127849936 | 0 |
| T CM | RPL11 | 0 | 1.099228859 | 0 |
| T EM | PFN1 | 0 | 0.986687362 | 0 |
| T EM | IL32 | 0 | 1.610342145 | 0 |
| T EM | CD3E | 1.3E-304 | 1.308663845 | 9.47E-301 |
| T EM | CD2 | 1.1E-287 | 1.467312932 | 6.15E-284 |
| T EM | LCK | 2.6E-272 | 1.262953758 | 1.17E-268 |
| T EM | RPSA | 4.4E-253 | 0.814624369 | 1.66E-249 |
| T EM | CALM1 | 7.9E-253 | 0.715068698 | 2.52E-249 |
| T EM | ARHGDIB | 4.6E-251 | 0.718464315 | 1.29E-247 |
| T EM | CNN2 | 1.9E-232 | 1.090877652 | 4.77E-229 |
| T EM | PPDPF | 1.9E-221 | 0.780041814 | 4.36E-218 |
| T EM | HMGB1 | 2.1E-220 | 0.805478156 | 4.24E-217 |
| T EM | C12orf75 | 9.5E-216 | 1.496819973 | 1.78E-212 |
| T EM | GZMA | 1.5E-208 | 1.859913707 | 2.62E-205 |
| T EM | CFL1 | 2.8E-208 | 0.597434163 | 4.46E-205 |
| T EM | GAPDH | 8.5E-204 | 0.773029089 | 1.28E-200 |
| T EM | HNRNPA1 | 2E-199 | 0.635649562 | 2.79E-196 |
| T EM | CD3D | 1.6E-196 | 1.050539613 | 2.15E-193 |
| T EM | HSPA8 | 8.1E-196 | 0.768334746 | 1.01E-192 |
| T EM | PPIA | 1.4E-193 | 0.677782595 | 1.6E-190 |
| T EM | CORO1A | 3.3E-193 | 0.678872168 | 3.75E-190 |
| T EM | TMSB4X | 3E-192 | 0.701423883 | 3.18E-189 |
| T EM | RPLP0 | 5.7E-191 | 0.735651016 | 5.82E-188 |
| T EM | NPM1 | 4.2E-189 | 0.708239377 | 4.14E-186 |
| T EM | CD3G | 2.1E-188 | 1.052509427 | 1.94E-185 |
| T EM | RPL35 | 3.8E-183 | 0.589234889 | 3.32E-180 |
| T Reg | TRAC | 2.1E-186 | 2.504023075 | 4.74E-182 |
| T Reg | CD2 | 9.6E-168 | 2.014061689 | 1.08E-163 |
| T Reg | LCK | 9.8E-150 | 1.721353292 | 7.33E-146 |
| T Reg | BCL11B | 1.9E-147 | 2.586610556 | 1.05E-143 |
| T Reg | SPOCK2 | 1.4E-142 | 1.985653996 | 6.27E-139 |
| T Reg | PRDX2 | 1.2E-141 | 1.983315706 | 4.59E-138 |
| T Reg | ACAP1 | 2.4E-138 | 1.722847581 | 7.82E-135 |

| | | | | |
|---|---|---|---|---|
| T Reg | CD3G | 5.5E-135 | 1.680864692 | 1.54E-131 |
| T Reg | TRAF3IP3 | 2.2E-134 | 1.42104578 | 5.38E-131 |
| T Reg | RCAN3 | 1.1E-121 | 2.344667435 | 2.41E-118 |
| T Reg | NOSIP | 1.2E-116 | 1.687200785 | 2.38E-113 |
| T Reg | LINC00861 | 1.6E-114 | 2.534571886 | 2.99E-111 |
| T Reg | ETS1 | 8.6E-107 | 1.445593357 | 1.48E-103 |
| T Reg | LINC-PINT | 2.1E-106 | 2.737786531 | 3.31E-103 |
| T Reg | CD48 | 2.7E-106 | 1.103573203 | 4.1E-103 |
| T Reg | AAK1 | 7.8E-106 | 1.541362524 | 1.09E-102 |
| T Reg | LDHB | 4.4E-103 | 1.221625805 | 5.75E-100 |
| T Reg | SEPT1 | 7.4E-103 | 1.73498702 | 9.17E-100 |
| T Reg | RPS20 | 3.5E-102 | 0.99298352 | 4.13E-99 |
| T Reg | BIN1 | 8E-102 | 1.613268495 | 8.99E-99 |
| T Reg | MZT2B | 2.9E-101 | 1.265387058 | 3.115E-98 |
| T Reg | LEPROTL1 | 1.15E-97 | 1.378185511 | 1.178E-94 |
| T Reg | SEPT6 | 7.29E-96 | 1.290680408 | 7.112E-93 |
| T Reg | PIK3IP1 | 1.29E-95 | 1.614526391 | 1.203E-92 |
| T Reg | EVL | 3.22E-95 | 1.193714261 | 2.886E-92 |
| Tn | CD7 | 2.4E-145 | 1.697336078 | 8.8E-142 |
| Tn | ETS1 | 6.76E-92 | 1.095367551 | 1.011E-88 |
| Tn | PIK3IP1 | 1.82E-81 | 1.30270195 | 1.948E-78 |
| Tn | LINC00861 | 6.47E-80 | 1.70900321 | 6.601E-77 |
| Tn | TXK | 1.56E-77 | 2.393122911 | 1.524E-74 |
| Tn | OXNAD1 | 9.05E-76 | 2.057674646 | 8.458E-73 |
| Tn | TXNIP | 2.58E-75 | 0.686633885 | 2.319E-72 |
| Tn | CXCR4 | 2.02E-70 | 0.973678648 | 1.679E-67 |
| Tn | CD69 | 8.34E-68 | 0.95116365 | 6.451E-65 |
| Tn | RPS20 | 1.69E-66 | 0.62431258 | 1.266E-63 |
| Tn | SLC38A1 | 2.29E-66 | 1.239191532 | 1.659E-63 |
| Tn | STK17A | 1.48E-63 | 1.099060893 | 9.793E-61 |
| Tn | PDCD4 | 7.3E-62 | 1.224786997 | 4.552E-59 |
| Tn | ACAP1 | 2.05E-60 | 0.978610933 | 1.244E-57 |
| Tn | CD2 | 8.5E-58 | 1.040145159 | 4.653E-55 |
| Tn | FCMR | 3.39E-56 | 1.117510676 | 1.812E-53 |
| Tn | GCC2 | 3.5E-53 | 1.097834826 | 1.634E-50 |
| Tn | ATM | 2.73E-52 | 1.01020813 | 1.201E-49 |
| Tn | NOSIP | 7.53E-52 | 0.947219908 | 3.189E-49 |
| Tn | RPL27A | 1.15E-51 | 0.482233763 | 4.596E-49 |
| Tn | SNHG8 | 2.9E-51 | 0.827898383 | 1.14E-48 |
| Tn | PSIP1 | 6.3E-50 | 1.007369518 | 2.318E-47 |
| Tn | PNISR | 6.88E-50 | 0.649157584 | 2.488E-47 |

| | | | | |
|---|---|---|---|---|
| Tn | NPM1 | 2.42E-49 | 0.534767032 | 8.495E-47 |
| Tn | TOMM7 | 2.7E-49 | 0.523479581 | 9.325E-47 |
| cDC2 | CD74 | 2.7E-207 | 5.057557583 | 6.09E-203 |
| cDC2 | HLA-DRA | 2.5E-193 | 4.824946404 | 2.79E-189 |
| cDC2 | HLA-DRB1 | 2.6E-189 | 4.507218361 | 1.96E-185 |
| cDC2 | HLA-DPA1 | 9.1E-185 | 4.348133564 | 5.09E-181 |
| cDC2 | HLA-DPB1 | 5.8E-183 | 4.203994751 | 2.6E-179 |
| cDC2 | CST3 | 1.3E-174 | 4.506518364 | 4.92E-171 |
| cDC2 | HLA-DRB5 | 2.5E-161 | 4.534143925 | 7.95E-158 |
| cDC2 | HLA-DMA | 3.5E-156 | 3.360064745 | 9.95E-153 |
| cDC2 | HLA-DQA1 | 4.9E-154 | 4.728191853 | 1.21E-150 |
| cDC2 | HLA-DQB1 | 1.9E-140 | 4.238793373 | 4.31E-137 |
| cDC2 | FTH1 | 1E-131 | 1.777403235 | 2.03E-128 |
| cDC2 | PLD4 | 9E-124 | 6.3570261 | 1.68E-120 |
| cDC2 | GSTP1 | 1.4E-121 | 2.289432287 | 2.39E-118 |
| cDC2 | FCER1G | 3.3E-109 | 2.372119427 | 5.26E-106 |
| cDC2 | GABARAP | 4.6E-105 | 1.864873886 | 6.89E-102 |
| cDC2 | TYROBP | 5.9E-105 | 2.443982363 | 8.23E-102 |
| cDC2 | CYBA | 1.3E-100 | 1.42224133 | 1.757E-97 |
| cDC2 | GRN | 2.63E-94 | 2.188431263 | 3.282E-91 |
| cDC2 | GSN | 1.12E-89 | 2.682803154 | 1.317E-86 |
| cDC2 | FTL | 2.74E-85 | 1.364216805 | 2.954E-82 |
| cDC2 | RPLP0 | 2.77E-85 | 1.221267819 | 2.954E-82 |
| cDC2 | HLA-DMB | 3.07E-84 | 2.364899874 | 3.127E-81 |
| cDC2 | FCER1A | 2.49E-82 | 8.013721466 | 2.424E-79 |
| cDC2 | LSP1 | 7.61E-82 | 1.397130966 | 7.112E-79 |
| cDC2 | CAPG | 4.76E-81 | 1.980050206 | 4.268E-78 |
| pDC | PRSS57 | 6.59E-57 | 8.385089874 | 3.694E-53 |
| pDC | SOX4 | 1.53E-55 | 4.120506287 | 4.3E-52 |
| pDC | CDK6 | 3.4E-55 | 4.977421761 | 8.472E-52 |
| pDC | LAPTM4B | 4.31E-50 | 7.235749722 | 8.8E-47 |
| pDC | EBPL | 4.74E-50 | 3.683744431 | 8.866E-47 |
| pDC | SMIM24 | 1.04E-48 | 10.21585178 | 1.789E-45 |
| pDC | SNHG7 | 1.55E-48 | 2.896008968 | 2.479E-45 |
| pDC | STMN1 | 6.19E-48 | 3.380157232 | 8.244E-45 |
| pDC | TSC22D1 | 6.25E-48 | 4.35357666 | 8.244E-45 |
| pDC | ANKRD28 | 4.03E-47 | 3.901785135 | 4.764E-44 |
| pDC | AMD1 | 3.37E-46 | 2.746589661 | 3.775E-43 |
| pDC | NUCB2 | 4.16E-46 | 3.128644466 | 4.449E-43 |
| pDC | IMPDH2 | 5.53E-46 | 3.348272324 | 5.636E-43 |
| pDC | ZNRF1 | 1.15E-44 | 4.666395187 | 1.075E-41 |

| | | | | |
|---|---|---|---|---|
| pDC | HMGA1 | 3.85E-44 | 2.729388952 | 3.318E-41 |
| pDC | DDAH2 | 6.83E-44 | 3.38551259 | 5.675E-41 |
| pDC | SSBP2 | 7.58E-44 | 3.770020962 | 6E-41 |
| pDC | SPINT2 | 2.12E-42 | 2.846805096 | 1.535E-39 |
| pDC | BEX4 | 8.66E-42 | 3.606184006 | 6.071E-39 |
| pDC | EGFL7 | 3.82E-41 | 7.544454575 | 2.523E-38 |
| pDC | BEX2 | 1.28E-40 | 4.042181492 | 7.962E-38 |
| pDC | APEX1 | 3.83E-40 | 2.435176611 | 2.322E-37 |
| pDC | BZW2 | 7.5E-40 | 3.644649267 | 4.426E-37 |
| pDC | KDM5B | 8.14E-40 | 3.443030596 | 4.68E-37 |
| pDC | MYB | 1.57E-39 | 6.873373032 | 8.539E-37 |

**Table S3. Differentially expressed genes in each cell cluster**

The top 25 differentially expressed genes (DEGs) per cluster were ranked by $log_2$ fold change. DEGs with an adjusted p-value<0.05 using the Wilcoxon Rank Sum Test with Bonferroni correction were selected. *Acronyms: CD8+ T, cytotoxic CD8+ T cells; Class Mono, classical monocytes; Int Mono, intermediate monocytes; DC, dendritic cells; MTC, malignant T cells; CM, central memory; E/EM, effector/memory; Reg, regulatory; MP, macrophage; ML Mono, megakaryocyte-like monocytes; B, B cells; NK, natural killer cells; NK-Like Mono, natura killer cell-like monocytes; NC Mono, nonclassical monocytes; Tn, T-naïve cells; cDC2, type 2 conventional dendritic cells; pDC, plasmacytoid dendritic cells.*

| gene | qval | log2fc | mean |
|---|---|---|---|
| HACD1 | 0 | 2.509863 | 0.113629 |
| PLS3 | 0 | 2.44166 | 0.095377 |
| CD70 | 0 | 2.177189 | 0.122666 |
| NEDD4L | 0 | 1.94043 | 0.233707 |
| KIR3DL2 | 0 | 1.886088 | 0.103778 |
| TPO | 0 | 1.767456 | 0.096776 |
| IGFBP4 | 0 | 1.750654 | 0.202557 |
| PTHLH | 0 | 1.739237 | 0.071008 |
| LAIR2 | 0 | 1.727247 | 0.073169 |
| COL6A2 | 0 | 1.640846 | 0.107552 |
| CXCL13 | 0 | 1.609768 | 0.098866 |
| DUSP4 | 0 | 1.56176 | 0.108303 |
| AIRE | 0 | 1.49677 | 0.520241 |
| KLHL42 | 0 | 1.486158 | 0.137819 |
| CHN1 | 0 | 1.457556 | 0.059486 |
| LMNA | 0 | 1.456174 | 0.296649 |
| GNG4 | 0 | 1.448043 | 0.073369 |
| PDLIM1 | 0 | 1.425236 | 0.233056 |
| PTMS | 0 | 1.412676 | 0.179403 |
| TUSC3 | 0 | 1.40849 | 0.110564 |
| TSPAN2 | 0 | 1.402891 | 0.160644 |
| DNM3OS | 0 | 1.389316 | 0.081069 |
| CCR4 | 0 | 1.3837 | 0.166911 |
| CCR10 | 0 | 1.380784 | 0.07106 |
| CX3CR1 | 0 | 1.327656 | 0.081138 |
| PCED1B | 0 | -2.2466 | 0.051238 |
| ITGA4 | 0 | -2.01783 | 0.066787 |
| PLAC8 | 0 | -1.86594 | 0.061499 |
| SATB1 | 0 | -1.86033 | 0.068197 |
| CD7 | 0 | -1.55611 | 0.243773 |
| LINC00861 | 0 | -1.5198 | 0.142767 |
| OXNAD1 | 0 | -1.49084 | 0.105907 |
| C1orf162 | 0 | -1.4568 | 0.089908 |
| TRABD2A | 0 | -1.44754 | 0.054948 |
| NUCB2 | 0 | -1.33776 | 0.072825 |
| GSTP1 | 0 | -1.28612 | 0.078229 |
| UPP1 | 0 | -1.21085 | 0.054063 |
| ABLIM1 | 0 | -1.15278 | 0.114434 |

| | | | |
|---|---|---|---|
| PRDX2 | 0 | -1.14054 | 0.140578 |
| CHCHD10 | 0 | -1.12019 | 0.112606 |
| CISH | 0 | -1.09292 | 0.104363 |
| LINC01550 | 0 | -1.07082 | 0.055127 |
| IFI44L | 0 | -1.06992 | 0.051247 |
| PDCD4 | 0 | -0.99829 | 0.119053 |
| CD96 | 0 | -0.98633 | 0.091229 |
| APBB1IP | 0 | -0.9563 | 0.119896 |
| FOXP1 | 0 | -0.93934 | 0.173263 |
| KLRB1 | 0 | -0.88041 | 0.13266 |
| FHIT | 0 | -0.87461 | 0.059252 |
| XBP1 | 0 | -0.87376 | 0.118017 |

**Table S4. Differentially expressed genes in MTC CM vs. T CM**

The top 25 differentially upregulated and downregulated differentially expressed genes (DEGs) in malignant central memory T cells (MTC CM) compared to benign central memory T cells (T CM) in cutaneous T cell lymphoma (CTCL) patients were ranked by log2 fold change. DEGs with an adjusted p-value (q-value) <0.05, corrected for multiple testing using the Benjamini-Hochberg (FDR) procedure, were selected. The table includes the following columns: q-value (FDR-adjusted p-value), log2fc (log2 fold change), and mean (average expression).

| gene | qval | log2fc | mean |
|---|---|---|---|
| HSPA6 | 0 | 4.114767 | 0.14161 |
| RASD1 | 0 | 3.239503 | 0.059424 |
| NDUFA4L2 | 0 | 3.076283 | 0.062238 |
| NR4A1 | 0 | 2.945409 | 0.234997 |
| EGR1 | 0 | 2.944622 | 0.264574 |
| PLK2 | 0 | 2.845227 | 0.121721 |
| HSPA1B | 0 | 2.734522 | 0.305756 |
| HSPA2 | 0 | 2.727889 | 0.101437 |
| IER5L | 0 | 2.6568 | 0.051105 |
| PLCG2 | 0 | 2.648124 | 0.11248 |
| ATF3 | 0 | 2.518105 | 0.17083 |
| RASGEF1B | 0 | 2.409381 | 0.055284 |
| IER3 | 0 | 2.398943 | 0.16462 |
| FOSB | 0 | 2.391358 | 0.479218 |
| DNAJA4 | 0 | 2.381732 | 0.071297 |
| KCNQ1OT1 | 0 | 2.346339 | 0.137821 |
| HEXIM1 | 0 | 2.290243 | 0.336011 |
| TNFSF9 | 0 | 2.278614 | 0.080464 |
| GEM | 0 | 2.273945 | 0.079405 |
| AC007952.4 | 0 | 2.263094 | 0.1078 |
| HMOX1 | 0 | 2.226942 | 0.198073 |
| MAFF | 0 | 2.220741 | 0.075213 |
| CD83 | 0 | 2.126164 | 0.214394 |
| NR4A3 | 0 | 2.123712 | 0.141404 |
| ZNF331 | 0 | 2.106729 | 0.281252 |
| TYMS | 0 | -4.71252 | 0.125722 |
| RRM2 | 0 | -4.66424 | 0.114113 |
| UBE2C | 0 | -4.60012 | 0.079727 |
| BIRC5 | 0 | -3.71666 | 0.077269 |
| MKI67 | 0 | -3.51605 | 0.093071 |
| HIST1H1B | 0 | -3.46462 | 0.051442 |
| TK1 | 0 | -3.2547 | 0.06892 |
| TOP2A | 0 | -3.11743 | 0.115633 |
| CDT1 | 0 | -2.97545 | 0.053438 |
| TPX2 | 0 | -2.48347 | 0.056542 |
| MAD2L1 | 0 | -2.37071 | 0.060912 |
| TMEM256-PLSCR3 | 0 | -2.36854 | 0.075202 |
| CCR10 | 0 | -2.26829 | 0.115026 |
| DHFR | 0 | -2.22662 | 0.061298 |

| | | | |
|---|---|---|---|
| ASF1B | 0 | -2.13534 | 0.052278 |
| FUT7 | 0 | -2.07425 | 0.063562 |
| JAKMIP1 | 0 | -2.05731 | 0.061764 |
| MYCBP | 0 | -2.02889 | 0.066788 |
| CDCA7 | 0 | -1.97567 | 0.115203 |
| EEF1G | 0 | -1.91838 | 0.414177 |
| C11orf98 | 0 | -1.90265 | 0.093952 |
| SMCO4 | 0 | -1.83225 | 0.052236 |
| DTYMK | 0 | -1.73772 | 0.06692 |
| RFC4 | 0 | -1.70636 | 0.051465 |
| C12orf75 | 0 | -1.70115 | 0.290426 |

**Table S5. Differentially expressed genes in MTC E/EM vs. T EM**

The top 25 differentially upregulated and downregulated differentially expressed genes (DEGs) in malignant effector and effector memory T cells (MTC E/EM) compared to benign effector memory T cells (T EM) in cutaneous T cell lymphoma (CTCL) patients were ranked by log2 fold change. DEGs with an adjusted p-value (q-value) <0.05, corrected for multiple testing using the Benjamini-Hochberg (FDR) procedure, were selected. The table includes the following columns: q-value (FDR-adjusted p-value), log2fc (log2 fold change), and mean (average expression).

| gene | qval | log2fc | mean |
|---|---|---|---|
| CX3CR1 | 0 | 297.776 | 0.082452 |
| MLF1 | 0 | 297.776 | 0.130574 |
| TNFSF11 | 0 | 297.776 | 0.067457 |
| EGLN3 | 0 | 297.776 | 0.095328 |
| GEM | 0 | 297.776 | 0.135996 |
| KIR3DL2 | 0 | 297.776 | 0.111761 |
| DNM3OS | 0 | 297.776 | 0.08504 |
| EPCAM | 0 | 297.776 | 0.118209 |
| PLS3 | 0 | 297.776 | 0.284141 |
| HDAC9 | 0 | 297.776 | 0.064538 |
| TGFBR3 | 0.024745 | 3.309968 | 0.205909 |
| AC016831.7 | 0.017416 | 3.277908 | 0.221129 |
| CYSTM1 | 0.019446 | 2.924482 | 0.190796 |
| DUSP4 | 1.2E-05 | 2.81002 | 0.468909 |
| NR4A3 | 0.018509 | 2.754243 | 0.183999 |
| NEDD4L | 0.000919 | 2.745122 | 0.297949 |
| PTMS | 0.024745 | 2.466492 | 0.161381 |
| SLC5A3 | 0.012336 | 2.44824 | 0.188599 |
| CCR4 | 0.001048 | 2.370485 | 0.272235 |
| LMNA | 2.14E-12 | 2.364509 | 0.948709 |
| TSPAN2 | 0.01684 | 2.318517 | 0.173443 |
| IKZF2 | 0.007507 | 2.299237 | 0.203051 |
| CD70 | 0.018509 | 2.292651 | 0.168745 |
| ANKRD28 | 0.021699 | 2.255811 | 0.16163 |
| CDKN1A | 0.000973 | 2.214324 | 0.272257 |
| XAF1 | 0 | -1.80655 | 0.11718 |
| IFITM3 | 3.2E-12 | -1.66561 | 0.085332 |
| PLAC8 | 3.73E-08 | -1.59642 | 0.063804 |
| SATB1 | 4.3E-08 | -1.50993 | 0.072237 |
| GSTP1 | 9.4E-08 | -1.49162 | 0.071042 |
| IFI6 | 0 | -1.47653 | 0.20896 |
| UPP1 | 9.13E-06 | -1.45604 | 0.056114 |
| EIF2AK2 | 2.37E-09 | -1.38809 | 0.10316 |
| TRIM22 | 0 | -1.38411 | 0.175569 |
| TRABD2A | 6.94E-05 | -1.37237 | 0.054522 |
| OXNAD1 | 1.02E-09 | -1.35789 | 0.113105 |
| MX1 | 3.66E-12 | -1.3298 | 0.147565 |
| OAS1 | 1.11E-05 | -1.32775 | 0.069296 |
| OAS2 | 8.48E-06 | -1.31958 | 0.072425 |
| GBP1 | 5.47E-05 | -1.22266 | 0.074384 |

| | | | |
|---|---|---|---|
| LPAR6 | 0.000191 | -1.20021 | 0.068792 |
| CISH | 8.89E-06 | -1.19808 | 0.09196 |
| CD96 | 4.91E-07 | -1.18875 | 0.113766 |
| TMEM204 | 8.65E-05 | -1.18853 | 0.076806 |
| LDLRAP1 | 3.97E-08 | -1.13916 | 0.147392 |
| IRF7 | 1.97E-08 | -1.08946 | 0.17145 |
| LINC00861 | 5.24E-09 | -1.08895 | 0.183004 |
| GIMAP5 | 9.42E-05 | -1.08544 | 0.095496 |
| SAMD9L | 0.000918 | -1.07832 | 0.075482 |
| MX2 | 1E-05 | -1.05484 | 0.124984 |

**Table S6. Differentially expressed genes in MTC Reg vs. T Reg**

The top 25 differentially upregulated and downregulated differentially expressed genes (DEGs) in malignant regulatory T cells (MTC Reg) compared to benign regulatory T cells (T Reg) in cutaneous T cell lymphoma (CTCL) patients were ranked by log2 fold change. DEGs with an adjusted p-value (q-value) <0.05, corrected for multiple testing using the Benjamini-Hochberg (FDR) procedure, were selected. The table includes the following columns: q-value (FDR-adjusted p-value), log2fc (log2 fold change), and mean (average expression).

| gene | qval | log2fc | mean |
|---|---|---|---|
| PTPRCAP | 0 | 2.204648 | 0.297684 |
| PTP4A1 | 0 | 2.187884 | 0.120361 |
| HLA-DRB5 | 0 | 2.059033 | 0.922416 |
| IGKV2-30 | 0 | 1.930552 | 0.108779 |
| CD99 | 0 | 1.830564 | 0.203576 |
| IFI44L | 0 | 1.757731 | 0.213809 |
| AKAP17A | 0 | 1.722752 | 0.074434 |
| CDKN1A | 0 | 1.715687 | 0.08055 |
| AC245100.1 | 0 | 1.672851 | 0.09135 |
| SLC25A6 | 0 | 1.651423 | 0.625785 |
| IGHV4-34 | 0 | 1.629368 | 0.115847 |
| IGHV3-21 | 0 | 1.579901 | 0.158601 |
| ATP6V0C | 0 | 1.578001 | 0.26182 |
| P2RY8 | 0 | 1.540341 | 0.114065 |
| IFITM1 | 0 | 1.521891 | 0.487191 |
| EGR1 | 0 | 1.504525 | 0.055531 |
| IGHV3-48 | 0 | 1.500048 | 0.102058 |
| IGLV1-51 | 7.51E-08 | 1.483603 | 0.064132 |
| GTPBP6 | 0 | 1.389999 | 0.061811 |
| IGHV4-39 | 4.56E-12 | 1.379987 | 0.105376 |
| IGLV3-1 | 0 | 1.308325 | 0.1765 |
| COL6A3 | 3.97E-16 | 1.306146 | 0.054172 |
| NR4A1 | 0 | 1.269301 | 0.072652 |
| IFI6 | 0 | 1.265174 | 0.172231 |
| IGKV1-12 | 4.22E-14 | 1.22565 | 0.050584 |
| AC007950.1 | 0 | -283.913 | 0.193011 |
| AL645728.1 | 0 | -283.913 | 0.052531 |
| OSBPL10-AS1 | 2.82E-07 | -5.91548 | 0.087253 |
| ANGPTL1 | 1.24E-06 | -5.44709 | 0.056908 |
| SPON1 | 5.97E-08 | -5.28332 | 0.064837 |
| AC012368.1 | 1.64E-10 | -4.9578 | 0.075087 |
| GPR174 | 1.45E-12 | -4.84262 | 0.085654 |
| TIGD5 | 1.46E-12 | -4.82563 | 0.08586 |
| GPM6B | 1.28E-08 | -4.76959 | 0.052914 |
| AC129492.1 | 1.72E-09 | -4.63221 | 0.05413 |
| PF4 | 3.3E-11 | -4.47036 | 0.061867 |
| LRRC75A | 0 | -4.17371 | 0.10107 |
| U2AF1L5 | 1.49E-11 | -4.0962 | 0.05443 |
| GDF7 | 0 | -4.03266 | 0.087304 |
| CBWD3 | 0 | -4.02501 | 0.118899 |

| | | | |
|---|---|---|---|
| GPR82 | 1.65E-11 | -3.97542 | 0.051392 |
| ARL17B | 3.84E-14 | -3.89177 | 0.064146 |
| ANGPT2 | 1.54E-14 | -3.8695 | 0.062575 |
| SLC16A4 | 6.31E-14 | -3.82934 | 0.059069 |
| PLCE1 | 2.28E-12 | -3.81743 | 0.051384 |
| FLCN | 0 | -3.80843 | 0.081171 |
| FAM153C | 1.63E-12 | -3.78865 | 0.051906 |
| C1orf56 | 0 | -3.78207 | 0.268202 |
| HCG11 | 3.97E-16 | -3.77499 | 0.067916 |
| P2RX5-TAX1BP3 | 0 | -3.76127 | 0.102284 |

**Table S7. Differentially expressed genes in B Cells of CTCL patients vs. healthy controls**

The top 25 differentially upregulated and downregulated genes in B cells from patients with cutaneous T cell lymphoma (CTCL), including Sézary syndrome and leukemic mycosis fungoides, compared to healthy controls. Genes were ranked by log2 fold change. Differentially expressed genes (DEGs) with an adjusted p-value (q-value) <0.05, corrected for multiple testing using the Benjamini-Hochberg (FDR) procedure, were selected. The table includes the following columns: q-value (FDR-adjusted p-value), log2fc (log2 fold change), and mean (average expression).

| gene | qval | log2fc | mean |
|---|---|---|---|
| CD99 | 0 | 2.669226 | 0.298811 |
| CSF2RA | 0 | 2.600635 | 0.050871 |
| HLA-DRB5 | 0 | 2.584502 | 0.441027 |
| SCO2 | 0 | 2.498045 | 0.111132 |
| PTP4A1 | 0 | 2.482298 | 0.050776 |
| SLC25A6 | 0 | 2.456743 | 0.585514 |
| HLA-DQA2 | 0 | 2.315161 | 0.090049 |
| IL32 | 0 | 2.254126 | 0.102186 |
| DUSP2 | 0 | 2.092933 | 0.120795 |
| ATP6V0C | 0 | 2.063411 | 0.378556 |
| CCL4 | 0 | 2.02859 | 0.080501 |
| WDR83OS | 0 | 2.026992 | 0.291444 |
| NR4A1 | 0 | 1.973882 | 0.113203 |
| EEF1G | 0 | 1.899744 | 0.175541 |
| CD83 | 0 | 1.858036 | 0.13687 |
| NME2 | 0 | 1.841362 | 0.332602 |
| HLA-DQA1 | 0 | 1.821424 | 0.13639 |
| CCL3 | 0 | 1.8119 | 0.159412 |
| EGR2 | 0 | 1.698165 | 0.069615 |
| RBKS | 0 | 1.68838 | 0.056677 |
| IL1B | 0 | 1.659863 | 0.157601 |
| IFITM1 | 0 | 1.587796 | 0.270852 |
| ATF3 | 0 | 1.509998 | 0.064576 |
| NBPF19 | 0 | 1.479769 | 0.102932 |
| A1BG | 0 | 1.457523 | 0.079872 |
| ALPL | 0 | -283.913 | 0.096514 |
| CD177 | 0 | -283.913 | 0.114852 |
| AC007950.1 | 0 | -8.83432 | 0.44669 |
| CA4 | 3.97E-07 | -7.91638 | 0.062152 |
| DAAM2 | 0 | -6.61694 | 0.168966 |
| CCNJL | 0 | -6.35411 | 0.057331 |
| ARG1 | 0 | -6.29043 | 0.10313 |
| AC129492.1 | 0 | -6.22105 | 0.282921 |
| ZDHHC19 | 0 | -5.99133 | 0.050716 |
| OLAH | 0 | -5.89941 | 0.087222 |
| AMPH | 0 | -5.79501 | 0.300673 |
| IL18RAP | 0 | -5.40586 | 0.071264 |
| IL18R1 | 0 | -5.12678 | 0.098658 |
| MME | 0 | -5.00785 | 0.092502 |
| ITGB3 | 0 | -4.99989 | 0.077409 |

| | | | |
|---|---|---|---|
| SH2D4A | 0 | -4.96199 | 0.055308 |
| ADGRG3 | 0 | -4.87806 | 0.082209 |
| ANXA3 | 0 | -4.85379 | 0.086163 |
| MAOA | 0 | -4.83089 | 0.084331 |
| IGHA1 | 0 | -4.55383 | 0.108679 |
| CXCR1 | 0 | -4.51858 | 0.087939 |
| ABLIM3 | 0 | -4.49781 | 0.06519 |
| PTCRA | 0 | -4.118 | 0.053426 |
| HPGD | 0 | -4.00535 | 0.112635 |
| IGKC | 0 | -3.99919 | 0.109335 |

**Table S8. Differentially expressed genes in monocytes of CTCL patients vs. healthy controls**

The top 25 differentially upregulated and downregulated genes in monocytes from cutaneous T cell lymphoma (CTCL) patients versus healthy controls. Genes were ranked by log2 fold change. DEGs with a q-value <0.05, adjusted using the Benjamini-Hochberg (FDR) method, were included. Columns presented are: q-value, log2fc, and mean expression.

| gene | qval | log2fc | mean |
|---|---|---|---|
| LAT | 3.57E-06 | 3.476533 | 0.24838 |
| IL32 | 0 | 2.790161 | 0.778548 |
| IL7R | 1.47E-09 | 2.776181 | 0.351861 |
| CD3D | 2.07E-10 | 2.532662 | 0.367445 |
| CD3E | 8.38E-13 | 2.45468 | 0.461573 |
| CD3G | 5.18E-07 | 2.184682 | 0.215879 |
| CD27 | 4.86E-06 | 1.919135 | 0.180572 |
| PTPRCAP | 5.09E-09 | 1.904221 | 0.302072 |
| CD5 | 0.000466 | 1.853396 | 0.106035 |
| SPOCK2 | 2.47E-05 | 1.793213 | 0.156652 |
| LCK | 1.61E-06 | 1.766795 | 0.205696 |
| AQP3 | 1.76E-06 | 1.734558 | 0.205805 |
| LEF1 | 4.76E-06 | 1.689604 | 0.190336 |
| IFITM1 | 0 | 1.553029 | 0.830668 |
| ID3 | 0.000295 | 1.528046 | 0.12561 |
| CDKN1A | 6.57E-06 | 1.497782 | 0.200072 |
| LIME1 | 4.38E-06 | 1.476242 | 0.2089 |
| CCR7 | 5.47E-06 | 1.403518 | 0.22014 |
| CRIP1 | 0 | 1.390393 | 0.737052 |
| P2RY8 | 0.000568 | 1.381606 | 0.12288 |
| TRIB1 | 0.011744 | 1.367376 | 0.066201 |
| LMNA | 2.5E-07 | 1.359333 | 0.29862 |
| LTB | 3.94E-14 | 1.306232 | 0.661298 |
| HLA-DQA2 | 4.56E-07 | 1.292167 | 0.312265 |
| SYNE2 | 0.000212 | 1.291037 | 0.150994 |
| PRSS57 | 0 | -5.34836 | 0.272078 |
| LAPTM4B | 0 | -4.19166 | 0.157068 |
| MYB | 0 | -3.82549 | 0.201094 |
| ZNRF1 | 0 | -3.32355 | 0.149307 |
| MAP7 | 0 | -3.3156 | 0.111349 |
| FTX | 4.43E-14 | -3.29558 | 0.095588 |
| EVA1B | 0 | -3.13655 | 0.103075 |
| SLC39A3 | 0 | -2.85301 | 0.149135 |
| ATP8B4 | 0 | -2.82859 | 0.124242 |
| MLLT3 | 0 | -2.81654 | 0.171118 |
| SNHG19 | 0 | -2.78746 | 0.105576 |
| FAM117A | 0 | -2.75997 | 0.167 |
| HIST3H2A | 0 | -2.68252 | 0.14264 |
| C11orf74 | 0 | -2.63974 | 0.101671 |
| PDZD8 | 0 | -2.55589 | 0.13589 |

| | | | |
|---|---|---|---|
| BEX2 | 0 | -2.51053 | 0.145012 |
| GOLIM4 | 1.48E-14 | -2.48067 | 0.097181 |
| CASP6 | 7.88E-15 | -2.47729 | 0.100462 |
| GCSH | 1.55E-12 | -2.47149 | 0.081898 |
| AGPS | 0 | -2.46832 | 0.149517 |
| GSE1 | 0 | -2.46601 | 0.103186 |
| FHL1 | 7.88E-15 | -2.46499 | 0.100658 |
| SAP30 | 1.74E-11 | -2.39076 | 0.075013 |
| CLEC11A | 0 | -2.39035 | 0.118837 |
| PRKAR2B | 0 | -2.37612 | 0.109186 |

**Table S9. Differentially expressed genes in dendritic cells of CTCL patients vs. healthy controls**

The top 25 differentially upregulated and downregulated DEGs in dendritic cells from cutaneous T cell lymphoma (CTCL) patients versus healthy controls, ranked by log2 fold change. DEGs were selected based on an FDR-adjusted q-value <0.05 using the Benjamini-Hochberg correction. Table columns include: q-value, log2fc, and mean expression.

| gene | qval | log2fc | mean |
|---|---|---|---|
| HLA-DRB5 | 0 | 2.879126 | 0.096005 |
| PTP4A1 | 0 | 2.677696 | 0.091731 |
| PTPRCAP | 0 | 2.556597 | 0.379825 |
| CD99 | 0 | 2.075526 | 0.719643 |
| SLC25A6 | 0 | 2.041345 | 0.501535 |
| IFI44L | 0 | 2.030567 | 0.09364 |
| COL6A3 | 0 | 1.935815 | 0.06132 |
| DIABLO | 0 | 1.917681 | 0.050406 |
| VAMP7 | 0 | 1.876816 | 0.050774 |
| P2RY8 | 0 | 1.867525 | 0.132974 |
| NME2 | 0 | 1.816237 | 0.250667 |
| GTPBP6 | 0 | 1.815905 | 0.080447 |
| NBPF19 | 0 | 1.804227 | 0.058873 |
| AKAP17A | 0 | 1.794085 | 0.076033 |
| EEF1G | 0 | 1.793207 | 0.135608 |
| AC245100.1 | 0 | 1.71499 | 0.050606 |
| ATP6V0C | 0 | 1.710443 | 0.266961 |
| OVCA2 | 0 | 1.59904 | 0.072641 |
| WDR83OS | 0 | 1.559669 | 0.335635 |
| LIME1 | 0 | 1.550847 | 0.124924 |
| HLA-DRB1 | 0 | 1.528766 | 0.233903 |
| KIR2DL3 | 0 | 1.402651 | 0.122237 |
| ALDOA | 0 | 1.278934 | 0.560829 |
| ZC3H11A | 0 | 1.243875 | 0.085019 |
| LMNA | 0 | 1.240459 | 0.065714 |
| TREML1 | 0 | -283.913 | 0.104837 |
| AC007950.1 | 0 | -283.913 | 0.137626 |
| ITGB3 | 0 | -283.913 | 0.058841 |
| ITGA2B | 0 | -6.4419 | 0.118951 |
| GP9 | 2.75E-14 | -5.88716 | 0.06179 |
| PF4V1 | 2.35E-15 | -5.85574 | 0.065876 |
| TUBB1 | 0 | -5.83672 | 0.116852 |
| F13A1 | 5.96E-16 | -5.44667 | 0.051787 |
| NRGN | 0 | -5.18433 | 0.237469 |
| PF4 | 0 | -5.07119 | 0.159652 |
| PPBP | 0 | -4.93258 | 0.274671 |
| PRKAR2B | 0 | -4.64184 | 0.092226 |
| SPARC | 0 | -4.4615 | 0.054347 |

| | | | |
|---|---|---|---|
| GNG11 | 0 | -4.3465 | 0.05954 |
| IGHA1 | 0 | -4.11452 | 0.150022 |
| AC129492.1 | 0 | -4.07712 | 0.075484 |
| IGHM | 0 | -4.03221 | 0.061106 |
| IGKC | 0 | -3.79347 | 0.153506 |
| IGFBP2 | 0 | -3.78881 | 0.07984 |
| AL645728.1 | 0 | -3.77998 | 0.062771 |
| PCDHGB6 | 0 | -3.59213 | 0.050939 |
| DOK6 | 0 | -3.58051 | 0.052386 |
| TSIX | 0 | -3.38433 | 0.07231 |
| MIA2 | 0 | -3.29841 | 0.166472 |
| HIPK1-AS1 | 0 | -3.26339 | 0.072775 |

**Table S10. Differentially expressed genes in NK cells of CTCL patients vs. healthy controls**

The top 25 differentially upregulated and downregulated genes in natural killer (NK) cells from cutaneous T cell lymphoma (CTCL) patients compared to healthy controls. Genes were ranked by log2 fold change. Only DEGs with a q-value <0.05 (FDR-adjusted using the Benjamini-Hochberg method) were included. Columns shown are: q-value, log2fc, and mean expression.
