## Supplementary Methods for "Single-Cell Profiling Reveals Targetable Malignant T-Cell Subtypes and Immune Evasion Pathways in Sézary Syndrome"

**Appendix S1: Methods**

**Patient Recruitment**

This study includes a cohort of 22 patients. From published reports, we integrate 12 patients with Sézary syndrome (SS) and 2 patients with mycosis fungoides (MF) with leukemic involvement, in addition to 8 newly recruited SS patients from UT Southwestern Medical Center. For some analyses, healthy controls were incorporated from publicly available datasets, including GSE171811 (n=1) and the Tabula Sapiens blood dataset (n=6).

Patients at UT Southwestern were recruited from the outpatient clinic of the Department of Dermatology. Inclusion criteria required a confirmed diagnosis of Sézary syndrome or mycosis fungoides with peripheral blood involvement, based on immunophenotypic (IP) flow cytometry and clinical evaluation by a dermatologist with expertise in CTCL. Demographic information, including age (range: 39–89 years) and sex distribution, is detailed in Table 1. Among patients from UT Southwestern, the mean percentage of Sézary cells in peripheral blood was 37%.

To provide a high-resolution characterization of Sézary syndrome, we integrated newly obtained samples with publicly available datasets. A comprehensive PubMed search identified four publicly available CTCL datasets in the Gene Expression Omnibus (GEO), which were harmonized with our dataset using stringent cross-study integration protocols (GSE124899, GSE146586, GSE165623, GSE171811).[1-4] Notably, samples from GSE124899 and GSE146586 included only CD45+ cells, and the sample from GSE165623 included sorted CD3+CD4+ helper T-cells vs remaining CD45+ cells in a 1:2 ratio.[1,2,3]

**Sample Collection, Processing, and Droplet-Based scRNA-seq**

Peripheral blood mononuclear cells (PBMCs) were isolated via Ficoll separation. Single-cell suspensions were subjected to processing prior to sequencing by using the Chromium Controller and Single-Cell 5' Library & Gel Bead Kits v1.1 and v2 (10X Genomics, Pleasanton, Calif), following manufacturer recommendations. For a subset of samples (Pool1, Pool2, and Pt_1, n=8), we integrated Cellular Indexing of Transcriptomes and Epitopes by Sequencing (CITE-seq) and flow cytometry to link transcriptomic data with protein-level expression, enabling robust surface marker analysis in tandem with RNA data. T-cell receptor (TCR) sequencing was performed on 15 publicly available samples, allowing clonal tracing and immune profiling. Details regarding the sequencing methodologies for these publicly available samples can be accessed directly from their respective GEO entries.

**Data Analysis**

Raw sequencing output was processed using the 10X Genomics pipeline (Cell Ranger v5.0.1). Reads were aligned to the GRCh38 human reference genome, generating filtered UMI count matrices. TCR sequences were analyzed using the cellranger vdj pipeline, aligning reads to the vdj_GRCh38_alts_ensembl-5.0.0 reference and assigning clonotypes based on frequency. Sample size was determined by sample availability rather than a power calculation.

**Secondary Analysis**

scRNA-seq data were processed using the AnnData (ad) framework (v.0.10.3) and analyzed with Scanpy (sc) (v.1.9.6) and SCVI (scvi) (v.1.0.2) in Python (v.3.11.10). Initially, 101,710 cells from 22 CTCL patients and 42,611 cells from 7 healthy controls were identified. Cells with fewer than 200 detected genes were excluded.

### Quality Control and Doublet Removal

Quality control (QC) was conducted on a sample-by-sample basis to ensure robust data processing and minimize technical artifacts. The following QC metrics were computed using sc.pp.calculate_qc_metrics, including mitochondrial gene content, ribosomal gene content, and total UMI counts. The following cutoffs were applied:

- **Mitochondrial gene content**: <25%
- **Log1p total UMI counts**: within 5 standard deviations of the median
- **Log1p number of genes detected**: within 5 standard deviations of the median
- **Percentage of counts in top 20 genes**: within 5 standard deviations of the median
- **Percentage of ribosomal genes**: within 3 standard deviations of the median
- **Mitochondrial gene percentage**: <4 standard deviations from the median (after already removing cells with >25% mitochondrial gene content)

These QC parameters were selected to optimize data distribution and reduce technical noise while preserving biologically relevant cell populations. After QC refinement, adjustments were made on a per-sample basis, including dataset-specific modifications such as reducing log1p_n_genes_by_counts cutoffs to 4 standard deviations from the median for SS4Blood, SS5Blood, GSE124, and GSE146_3, and minor further adjustments for other datasets as needed.

Doublets were identified using scVI-based doublet detection. Cells were flagged as doublets if their doublet score exceeded the singlet score by >1 and if the scVI model predicted a cell was a doublet. This approach yielded an expected doublet rate consistent with the estimated number of captured cells per 10X Genomics guidelines. After initial processing, a small population of double-positive CD4+ and CD8+ T cells were presumed to represent doublets and removed from further analysis.

Following QC and doublet removal, 70,871 cells from CTCL patients and 37,100 from healthy controls remained.

### TCR Sequence Data and Clonotype Annotation

For cells with available T-cell receptor (TCR) sequencing data, clonotype information was appended to the metadata. TCR sequences were analyzed by grouping cells based on the relative frequencies of their CD3R amino acid sequences.

We assessed whether any samples shared identical CD3R amino acid sequences among their most clonally expanded T-cell populations, but no overlap was detected. To facilitate clonotype-based analyses, we introduced a metadata variable called clonotype_color, assigning the top 11 most expanded T-cell clonotypes as C1, C2, C3, ... C11, while all remaining, less frequent clonotypes were labeled as “NA.”

**Clustering and Dimensionality Reduction**

Clustering was performed using sc.pp.neighbors with n_neighbors=10 and n_pcs=30, followed by Leiden clustering with iterative resolution tuning to achieve optimal separation of biologically relevant populations. Cluster relationships were visualized using Uniform Manifold Approximation and Projection (UMAP).

To correct for batch effects and ensure consistent alignment across diverse sample types, we evaluated multiple integration strategies and selected Harmony, as it yielded the most stable and biologically coherent UMAP projections. Mitochondrial and ribosomal gene expression were regressed out to mitigate technical artifacts and improve clustering accuracy.

**Cell Annotation and Malignant Cell Identification**

Cell-type annotation was performed using a combination of automated and manual approaches. Initial predictions were generated using CellTypist (v.1.6.3), leveraging publicly available reference atlases such as Tabula Sapiens and the CellTypist’s “Immune_All_Low” high-resolution dataset. These automated annotations were refined manually based on established canonical markers from the literature to ensure high-fidelity cell-type identification.

Malignant T cells were identified using a multi-step approach. First, clonal density was assessed by overlaying TCR-labeled clones (e.g., C1, C2), and clusters where >50% of cells belonged to a dominant clone were classified as malignant, as consistent with prior literature.[1,2] Since TCR data were not available for all samples, additional validation was performed by comparing malignant and benign T cells against healthy controls, computing inferred copy number variations (CNVs) using infercnvpy (v.0.5.1.dev1+ge5943ea), and assessing expression patterns of key markers such as decreased CD7 and increased AIRE to confirm malignant cell classification. Subsets of malignant T cells were defined based on expression of canonical markers—*CCR7*, *SELL*, and *TCF7* in MTC CM; *CCR4*, *CD69*, and *GADD45B* in MTC E/EM; and *FOXP3*, *CTLA4*, and *IL12RA* in MTC Reg.

**Differential Expression Analysis**

Differential gene expression was analyzed using the Diffxpy (v.0.7.4) package. The top 25 upregulated and downregulated genes in malignant cells were identified relative to benign T cells overall and within CD4+ and CD8+ subsets. Genes with adjusted q-values < 0.05 and absolute log2 fold change > 0.5 were retained. To exclude confounding factors, genes starting with "TRBV", "TRAV", "TRGV", "TRDV", "RP", "MT", and "HB" (representing TCR, ribosomal, mitochondrial, and hemoglobin-associated genes) were removed. Genes with mean expression < 0.05 were also filtered out. Subsequent pathway analysis was conducted using gseapy, as outlined below, to identify dysregulated signaling networks relevant to CTCL pathogenesis.

**Gene Set Enrichment Analysis (GSEA)**

Gene Set Enrichment Analysis (GSEA) was conducted using the gseapy (v.1.0.3) package to uncover dysregulated biological processes and pathways in CTCL. Differentially expressed genes were ranked by log2 fold change and assessed for enrichment in hallmark and KEGG pathway gene sets. The gsea function was used with 10,000 permutations, applying the Benjamini-Hochberg correction for multiple comparisons. Pathways with adjusted p-values < 0.05 were considered significantly enriched. Cluster-specific pathway analysis was performed by separately analyzing malignant and benign T-cell subsets to identify distinct functional signatures. The results were visualized using enrichment plots, dot plots, and network representations to highlight key pathway alterations in CTCL.

**Cell-Cell Communication Analysis**

Cell-cell communication analysis was performed using the Liana (li) package (v.1.2.1), which integrates multiple ligand-receptor interaction inference methods into a consensus framework. To compare interactions between malignant and benign T cells, we applied li.mt.rank_aggregate.by_sample to all unpooled samples using the **consensus** model, running 10,000 permutations to identify significant interactions. Ligand-receptor pairs with **adjusted p-values <0.05** were considered significant.

To explore overarching communication themes, we employed **tensor decomposition** using li.multi.to_tensor_c2c. This allowed us to extract biologically relevant ligand-receptor interactions and analyze their functional implications. The **cell2cell pipeline** (c2c.analysis.run_tensor_cell2cell_pipeline) was used to assess cell-type-specific communication patterns and identify signaling pathways enriched in CTCL relative to healthy controls.

To compare T-cell interactions, we assessed malignant T cells relative to benign T cells, while interactions involving non-T cells were analyzed by comparing each population against its corresponding cell type in healthy controls. To further elucidate the functional consequences of cell-cell communication differences, we mapped ligand-receptor interactions to **PROGENy** pathways, identifying key signaling mechanisms underlying CTCL pathogenesis.


1. Borcherding N, Voigt AP, Liu V, Link BK et al. Single-Cell Profiling of Cutaneous T-Cell Lymphoma Reveals Underlying Heterogeneity Associated with Disease Progression. Clin Cancer Res 2019 May 15;25(10):2996-3005.
2. Borcherding N, Severson KJ, Henderson N, Ortolan LS et al. Single-cell analysis of Sézary syndrome reveals novel markers and shifting gene profiles associated with treatment. Blood Adv 2023 Feb 14;7(3):321-335
3. Rindler K, Bauer WM, Jonak C, Wielscher M, Shaw LE, Rojahn TB, Thaler FM, Porkert S, Simonitsch-Klupp I, Weninger W, Mayerhoefer ME, Farlik M, Brunner PM. Single-Cell RNA Sequencing Reveals Tissue Compartment-Specific Plasticity of Mycosis Fungoides Tumor Cells. Front Immunol. 2021 Apr 21;12:666935. doi: 10.3389/fimmu.2021.666935. PMID: 33968070; PMCID: PMC8097053
4. Herrera A, Cheng A, Mimitou EP, Seffens A, George D, Bar-Natan M, Heguy A, Ruggles KV, Scher JU, Hymes K, Latkowski JA, Ødum N, Kadin ME, Ouyang Z,

Geskin LJ, Smibert P, Buus TB, Koralov SB. Multimodal single-cell analysis of cutaneous T-cell lymphoma reveals distinct subclonal tissue-dependent signatures. Blood. 2021 Oct 21;138(16):1456-1464. doi: 10.1182/blood.2020009346. PMID: 34232982; PMCID: PMC8532199.
